# Accounting for contact tracing in epidemiological birth-death models

**DOI:** 10.1101/2024.09.09.24313296

**Authors:** Anna Zhukova, Olivier Gascuel

**Affiliations:** G5 Evolutionary Dynamics of Infectious Diseases, Institut Pasteur, Université de Paris, Paris, France; Bioinformatics and Biostatistics Hub, Institut Pasteur, Université de Paris, Paris, France; Institut de Systématique, Evolution, Biodiversité (ISYEB) - URM 7205 CNRS, Muséum National d’Histoire Naturelle, SU, EPHE & UA, Paris, France

## Abstract

Phylodynamics bridges the gap between classical epidemiology and pathogen genome sequence data by estimating epidemiological parameters from time-scaled pathogen phylogenetic trees. The models used in phylodynamics typically assume that the sampling procedure is independent between infected individuals. However, this assumption does not hold for many epidemics, in particular for such sexually transmitted infections as HIV-1, for which contact tracing schemes are included in health policies of many countries.

We extended phylodynamic multi-type birth-death (MTBD) models with contact tracing (CT), and developed a simulator to generate trees under MTBD and MTBD-CT models. We proposed a non-parametric test for detecting contact tracing in pathogen phylogenetic trees. Its application to simulated data showed that it is both highly specific and sensitive. For the simplest representative of the MTBD-CT family, the BD-CT(1) model, where only the last contact can be notified, we solved the differential equations and proposed a closed form solution for the likelihood function. We implemented a maximum-likelihood program, which estimates the BD-CT(1) model parameters and their confidence intervals from phylogenetic trees. It performed accurate parameter inference on BD and BD-CT(1) simulated data, and detected contact tracing in HIV-1 B epidemics in Zurich and the UK. Importantly, we showed that not accounting for contact tracing when it is present, leads to bias in parameter estimation with the BD model (overestimation of the becoming-non-infectious rate). This bias is also present, but greatly reduced, when the BD-CT(1) model is used on data where multiple contacts can be notified.

Our CT test, MTBD-CT tree simulator and BD-CT(1) parameter estimator are freely available at github.com/evolbioinfo/treesimulator and github.com/evolbioinfo/bdct.

**Author summary:** Phylodynamic models can estimate epidemiological parameters such as the number of secondary infections *R*_*e*_ from pathogen phylogenetic trees (i.e., genealogies, inferred from pathogen genomic sequences). These models do not account for contact tracing, and instead assume that detection of new cases is independent between infected individuals. However, especially for sexually transmitted infections, contact tracing plays an important role and is included in health policies of many countries.

We developed a phylodynamic model accounting for contact tracing and a test for the detection of contact tracing in pathogen phylogenetic trees. Our test and epidemiological parameter estimator showed good performance on simulated and real data. We detected the presence of contact tracing in the HIV-1 B epidemics in Zurich and the UK, and corrected the previous parameter estimates made without accounting for contact tracing.

## Introduction

The interaction of epidemiological and evolutionary processes leaves a footprint in pathogen genomes. Phylodynamics leverages this footprint to estimate epidemiological parameters [1, 2]. It relies on models that bridge the gap between traditional epidemiology and sequence data by estimating such parameters as the basic reproduction number, *R*_0_, from topology and branch lengths of pathogen phylogenies (i.e., genealogies of the pathogen population, approximating the transmission trees) combined with metadata on the samples. Classical epidemiological methods make inferences from incidence curves, which in the presence of imported cases might be biased and give appearance of elevated local transmission. In contrast, phylodynamic approach can overcome such biases as it uses the transmission structure as one of its information sources [3]. Using genomic data with phylodynamic estimations can provide valuable insights for preventing epidemic spread.

Phylodynamic models can be classified into two main families: coalescent [4–6] and birth-death (BD) [7–10]. Coalescent models are often preferred for estimating deterministic population dynamics, however, for highly stochastic processes, such as the dynamics of emerging pathogens, BD models are better adapted [11]. In the classic BD model with incomplete sampling [9], births represent pathogen transmission events (happening at a constant transmission rate), while deaths correspond to becoming non-infectious (e.g., due to healing, self-isolation, starting a treatment, or death, modeled with a constant removal rate).

However, the spread and detection of an epidemic can be non-homogeneous and depend on different factors, including governmental health policies (e.g., quarantine measures or development of pathogen detection tests), pathogen evolution over time (e.g., some SARS-CoV-2 variants are more transmissible than the others), differences between host individuals (e.g., due to their immune system particularities or to their behavior).

To allow for heterogeneity at the population level, the multi-type birth-death (MTBD) extension of the classical BD model was developed by Stadler *et al*. [12]. The MTBD framework allows for different types of individual states (and changes between them, modeled with state change rates). MTBD models are phylodynamic analogies of compartmental models in classical epidemiology (e.g., SIR, Susceptible-Infectious-Removed). Examples of MTBD models include the Birth-Death Exposed-Infectious (BDEI) model, which was designed for pathogens featuring an incubation period between the moments of infection and of becoming infectious (e.g., Ebola and SARS-CoV-2), and the Birth-Death with Super-Spreading model (BDSS) accounting for the fact that some individuals might spread the pathogen (e.g., HIV) more than the others [13].

To allow for heterogeneity over time, Stadler *et al*. [14] developed one-state Bayesian birth-death skyline plot (BDSKY) that divides the time into intervals and allows for different piecewise-constant rates on them. Kühnert *et al*. [15] combined the MTBD model with the BDSKY to allow for both piecewise-constant rate changes over time and multiple individual types. In particular the skyline approach can be useful to account for changes in the sampling policies (e.g., before/after HIV discovery or before/after PCR test spread for SARS-CoV-2).

Another important source of heterogeneity that none of these models accounts for is caused by non-independent sampling. One can set different sampling probabilities for different types of individuals in the MTBD framework, or change of the sampling probability between time intervals with the BDSKY, however, none of these models can account for correlation effects in sampling, in particular due to contact tracing (CT). CT (or partner notification, for sexually transmitted infections) is a process in which contacts of detected infected individuals are identified and offered testing for infection. It has been applied to various infectious diseases, including syphilis, HIV, tuberculosis, Ebola and COVID-19, and shown to successfully reduce transmission [16]. For instance, for HIV, partner notification showed increased early referral and initiation of treatment, and several countries include it in their national HIV testing services policies [17].

In this study we propose an extension of MTBD models that allows for modeling non-random sampling due to contact tracing, MTBD-CT. We describe the −CT extension and its assumptions, and present a transmission tree simulator under MTBD-CT models. We propose a non-parametric test for detecting contact tracing in pathogen phylogenetic trees. For the simplest representative of this model family, the BD-CT(1) model, where only the latest contact can be notified, we also derive the equations (and their closed-form solution) for its likelihood calculation and present a maximum-likelihood parameter estimator. We test our BD-CT(1) parameter estimator on data simulated under the the classical BD and the BD-CT models where one, two or all the contacts can be notified. We then apply it to the HIV-1 B epidemics in Zurich and in the UK. Importantly, we show that not accounting for contact tracing when it is present leads to an important bias in parameter estimation with the BD model. The BD-CT(1) parameter estimator performs well both in absence of contact tracing and in its presence when only the last contact can be notified. It shows some bias, however much less than BD, when multiple contacts can be notified.

## Materials and methods

In a pathogen transmission tree 𝒥 (approximated by a time-scaled pathogen phylogeny) the tips represent sampled pathogens while bifurcations (i.e., internal nodes) correspond to pathogen transmissions. The tree branches are measured in units of time, and the time goes forward, from the tree root (representing the beginning of the (sub-)epidemic, at time *t* = 0) to the time of the last sampled tip (at *t* = *T*).

### Extending Multi-Type Birth-Death models with Contact Tracing

#### Birth-Death (BD) model with incomplete sampling

Under the basic birth-death (BD) model [9] all the individuals are in the same state. They can transmit with a constant rate *λ*, get removed with a constant rate *ψ*, and their pathogen can be sampled upon removal with a constant probability *ρ*.

Under this model we can calculate the likelihood density *L*(𝒿| Θ) of a tree 𝒥under parameter values Θ = *{λ, ψ, ρ}* (see Mathematical formulation of BD and BD-CT(1) models). The likelihood density can then be used to estimate model parameters from a phylogenetic tree (e.g., by finding parameter values that maximize it, in the maximum-likelihood framework). In particular, it permits the estimation of the following epidemiological parameters:

- *effective reproduction number* 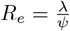, expected number of individuals directly infected by an infectious case;
- *infectious time* 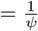, expected time during which an infectious individual can further spread the epidemic.

Note that the BD model is asymptotically unidentifiable (see Remark 3.4 in [9]). To become identifiable it requires one of its parameters to be fixed. In practice, it is often the sampling probability *ρ*, as it may be approximated from epidemiological data (e.g., the proportion of sampled cases among the declared ones) or the infectious time 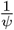 (estimated from observations of infected cases).

#### Multi-Type Birth-Death (MTBD) models

Multi-Type Birth-Death (MTBD) models [12] add population structure to the BD model by allowing different individual states, transmissions between them and state changes. A general MTBD model with *m* individual states has 2*m*^2^ + *m* parameters:

An individual in state *k ∈* {1, …, *m*} can be removed at a constant average rate *ψ*_*k*_ (with pathogen sampling probability *ρ*_*k*_), change their state to state *l ∈ {*1, …, *m}* at a constant average rate *µ*_*kl*_ (*k ?*= *l*), and transmit their pathogen to an individual in state *r∈ {*1, …, *m}* at a constant average rate *λ*_*kr*_. The time between events is hence modeled with exponential distributions.

#### Contact Tracing (CT) extension

We propose an extension of the MTBD models that adds contact tracing (CT). In the contact tracing process, an identified infected index patient notifies their potentially infected contacts (e.g., sexual or drug injection partners for HIV), either on their own (patient referral) or with the help of a health service provider (provider referral: a health adviser notifies the contacts without identifying the index patient; or contract referral: if the index patient fails to notify contacts within a specified time period, the health adviser proceeds to provider referral) [18]. The notified contacts are then provided with appropriate tests and treatment. Therefore, a successful detection of a new case B via contact tracing by an index patient A implies several conditions to be met: (i) A remembers the interaction with B and either notifies B on their own or gives the B’s details to a health advisor for assisted notification; (ii) B is successfully reached and notified; (iii) B agrees to take a test; (iv) B is indeed infected. For instance, according to the report from the Public Health England [19], in England in 2013 for HIV, reaching the stage (ii) had an estimated probability of about 50%; reaching the stage (iii) – of about 41% (as 83% of all the patients who attended a clinic through CT had a reported HIV test); and the stage (iv) – of less than 3%.

All these conditions are hardly identifiable separately from only sequence data, hence in our model we combine them into one, “an infected person B being successfully notified by A”, and model it with a constant notification probability *υ* which is applied upon sampling of A.

For simplicity, a positive draw with probability *υ* models a slightly modified scenario: (i) A remembers the interaction with B and either notifies B on their own or gives the B’s details to a health advisor for assisted notification; (ii’, similar to iv in the previous scenario) B is indeed infected, moreover, *the infection of either A or B is caused by the pathogen transmission during their interaction*; and either (iii’, same as ii) B is successfully reached and notified and (iv’, same as iii) agrees to take a test (which at the end of the considered time period might not yet be available in the database); or (iii”) *by the time of notification, B is already removed by other means* (already got sampled or stopped being infectious without sampling, e.g., died).

Note that in (ii’) in contrast to (iv) we exclude the possibility that B is indeed infected, however neither A infected B, nor B infected A (e.g., if they were both infected by a third person C). While in real life in such a situation B might still happen to get identified through tracing of A’s contacts, we consider such cases rare enough, and do not model them. From now on, we will use the term “contact” to denote a contact, during an interaction with whom a pathogen transmission happened.

Upon notification, if not yet removed, the contact’s pathogen is sampled almost instantaneously (modeled via a constant notified sampling rate *ϕ* >> *ψ*). Upon sampling, the contact might notify in their turn.

BD-CT hence has 3 classical BD and 2 additional parameters:

- *λ* – constant transmission rate: an infected individual transmits their pathogen to a newly infected individual with this rate. This leads to an internal node in the transmission tree;
- *ψ* – constant removal rate: an infected individual stops being infectious at this rate. This leads to a tip in the transmission tree (observed or hidden);
- *ρ* – constant sampling probability: upon removal the individual’s pathogen might get sampled with this probability. This leads to a tip in the transmission tree being observed;
- *υ* – constant contact tracing probability: upon sampling, a sampled individual’s contact might be notified with this probability. This leads to a removal rate change for the contact’s branch in the transmission tree;
- *ϕ* >> *ψ* – constant notified sampling rate: a notified contact (if not yet removed via the standard procedure) gets removed *and observed* with this rate. This leads to an observed tip in the transmission tree.

We add an additional hyper-parameter *κ*, which defines how many contacts a sampled individual might notify. Under the BD-CT(1) model (i.e., *κ* = 1) each sampled individual can notify only their last contact. Under the BD-CT(*κ*) model, each sampled individual can notify up to last *κ* contacts (their notification is independent, and depends for each of them on the probability *υ*). Note that *κ* = 0 gives a standard BD model with no contact tracing.

This model is a simplification of the contact tracing process, in particular it makes 3 assumptions:

1. only observed individuals can notify (instead of any removed individual); This is a realistic hypothesis in which an individual is “observed” because he or she enters a diagnostic and follow-up process that includes contact tracing. Without entering this process, he or she is not observed and does not notify.
2. successfully notified contacts get sampled (i.e., their pathogens are always observed upon removal, unlike the standard case where the observation happens with a sampling probability *ρ*); Realistically again, we assume here that the diagnostic and follow-up process works as expected, sampling notified contacts.
3. only the last *κ* contacts can get notified; In the case of sexually transmitted diseases, notifying only the last contact (*κ* = 1) could roughly correspond to notifying the “main” partner (e.g., spouse). Also note that the expected number of contacts per individual is *R*_*e*_, so depending on the pathogen, a low value of *κ* might be enough (e.g., 1 or 2 for HIV).

Note that the “last contact” relationship is not necessarily symmetrical. The last contact *j* of individual *i* might have transmitted their pathogen further (to individual *k*) after the interaction with *i*. Hence, while *j* is the last contact of *i, j*’s last contact is *k*, and, once notified, *j* might notify *k*.

The −CT(*κ*) extension can be generalized from BD to MTBD models: at the moment of sampling, an individual of any type can notify their last *κ* contacts (each independently, with the probability *υ*). If notified, the contact’s removal rate gets replaced by a constant sampling rate *ϕ >> ψ*_*k*_ *∀ k∈ {*1, …, *m}*. The −CT(*κ*) extension therefore adds two parameters to an MTBD model: *ϕ* and *υ*.

### MTBD-CT tree simulator and test datasets

We implemented a tree simulator that generates sampled transmission trees under MTBD and MTBD-CT(*κ*) models (with *m* states). It is Gillespie-based, generates state change, transmission and removal times, and only reconstructs the sampled parts of the tree to save memory and increase speed. We give its algorithmic details in S1 Appendix. The results in section “Testing for presence of Contact Tracing” show that the trees generated under MTBD and MTBD-CT(*κ*) models are different and easily distinguishable.

Using our simulator, we generated trees under three classical MTBD-family models: BD, BDEI (Birth-Death Exposed-Infectious [13]) and BDSS (Birth-Death with Super-Spreading [12]), and under their −CT versions: BD-CT(1), BDEI-CT(1), BDSS-CT(1), BD-CT(2), BDEI-CT(2), BDSS-CT(2), BD-CT(*∞*), BDEI-CT(*∞*) and BDSS-CT(*∞*) models. *κ* =*∞* here corresponds to the possibility to notify all the contacts of a sampled index case.

For each model, we generated 100 trees with 500–1 000 tips. For each tree the parameter values were drawn uniformly within the following bounds:

- 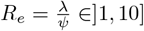,
- infectious time 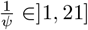,
- sampling probability *ρ ∈*]0.05, 0.75],
- *(for −CT models only)* ratio between the contact sampling rate after notification and the removal rate 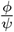 *∈*]10, 500],
- *(for −CT models only)* contact tracing probability *υ∈*]0.01*/ρ*, 0.75], (the lower bound was picked this way to ensure that the probability to be a notified contact is at least 0.01 = *ρυ*),
- *(for BDEI models only)* fraction of incubation period over total infection time 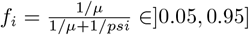,
- *(for BDSS models only)* fraction of superspreaders 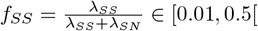,
- *(for BDSS models only)* super-spreading transmission ratio 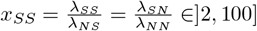.

We chose wide parameter ranges to cover a variety of epidemics. For instance, *R*_*e*_ *∈*]1, 10] covers epidemics such as HIV-1 (with *R*_*e*_ between 1 and 2 [20, 21]), influenza (with *R*_*e*_ also between 1 and 2 [22]), SARS-CoV-2 (with a potentially higher *R*_*e*_, e.g., between 2 and 4 for its Omicron variant [23]), and Ebola (with estimates between 1 and 8 [24]). The infectious time 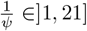 covers, on a yearly scale, such epidemics as HIV-1 B (whose average time of progression to AIDS in the absence of treatment is 10 years, and the time before treatment start is shorter). On a daily scale,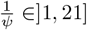 corresponds to]1 day - 3 weeks] and covers influenza (with infectious time of around 1 week), SARS-CoV-2 (up to 10 days since the symptom onset [25]) and Ebola (1-16 days [26]).

### Testing for presence of Contact Tracing

To access whether a given tree reveals contact tracing, we developed a non-parametric test based on cherries (two-tip clades).

The intuition behind the test is that in the presence of contact tracing the tree will contain more cherries whose tips are close in time. Indeed, in a transmission tree internal nodes correspond to transmissions while tips correspond to sampling events. Hence if a transmission tree is complete, a cherry corresponds to a transmission from an infected individual to their last contact, followed by sampling of both. If there is no contact tracing, these two sampling events are independent, while in the presence of contact tracing the first sampling might trigger the second one, and they will be close in time (e.g., the second cherry in Fig. 1a). For an incomplete transmission tree (where some infected individuals were not sampled), the difference would be that some cherries might include hidden transmissions. The contact tracing pattern should however still be observed for some of the cherries.

**Fig 1.**
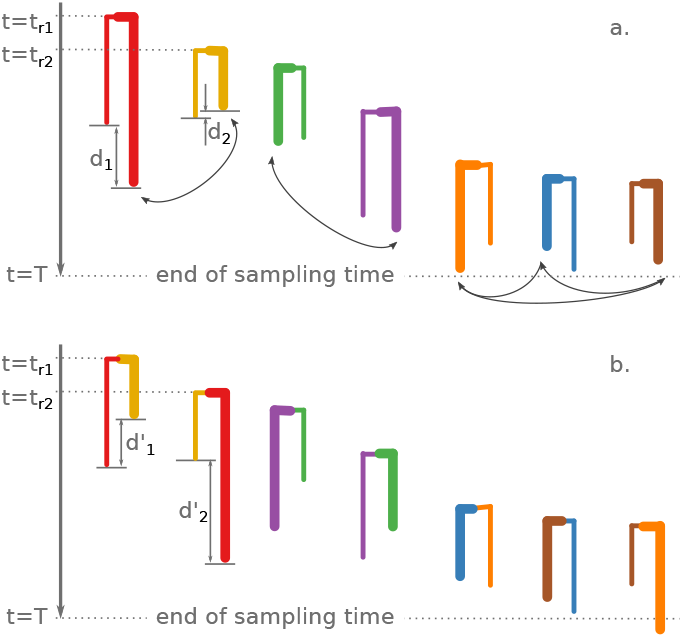
Reshuffled cherry generation. **a**. Cherries of the original tree sorted by their root times (times of the roots of the two oldest cherries, *t*_*r*1_ and *t*_*r*2_, are shown on the left). The randomly selected tip is shown with a bold branch for each cherry (e.g., the tip on the right for the leftmost cherry). The tip sampling time differences are shown for the two leftmost cherries (*d*_1_ and *d*_2_). **b**. Reshuffled cherries, which were obtained from the original ones by swapping the selected tips between the neighboring cherry couples (as shown with arrows in a.): the first and the second; the third and the forth. As the total number of cherries (7) is odd, the last three (instead of two) cherries exchanged their selected tips in a cycle. The reshuffled tip sampling time differences are shown for the two leftmost cherries (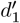 and 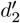).

More formally, in the absence of contact tracing (null hypothesis, H0), we assume that branch lengths of two cherry tips are independently drawn from the same distribution (which is the case for MTBD models). In the presence of contact tracing (alternative hypothesis, H1), we assume that for some of the cherries (ones that include notified contacts) their branch lengths are not independent and are similar. Therefore, in the absence of contact tracing two cherry branches taken randomly from the tree and two branches belonging to the same cherry should be indistinguishable, if they all start at about the same time. Being close in starting time is important as the tree is censored by the end of the sampling period: *t*≤ *T*. Hence the external branches that start close to the end of the sampling period would be on average shorter (as unless sampled before *T* they will stay hidden) than those that start closer to the root [27].

Let us assume that there are N cherries in the tree. For each cherry *i* (1 *≤ i ≤ N*) with tips *i*_0_ and *i*_1_, we calculate the difference between its tip times 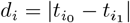. We then generate a collection of reshuffled cherries by (1) ordering the cherries by their root dates; (2) for each cherry *i* randomly selecting one of its tips *i*_*∗*_*∈*{*i*_*0*,_ *i*_*1*_}; (3) swapping the selected tips between the neighboring cherries: the tip 2*i*_*∗*_ gets swapped with the tip 2*i −* 1_*∗*_ (1 *≤i ≤N/*2) as illustrated in Fig. 1. (If the total number of cherries N is odd, instead of swapping the selected tips of the last two cherries, we swap the selected tips of the last three cherries in a cycle as in Fig. 1). We then calculate the time differences *d*_*i*_*′* between the tips of these reshuffled cherries. The test then compares *d*_*i*_ to *− d*_*i′*_ : *sign*(*d*_*i′*_ − *d*_*i*_) = *−* 1 if *d*_*i′*_ is larger than *d*_*i*_; = −1 if it is smaller; or = 0 if they are equal (this could happen as in real datasets dates are often truncated, e.g., to a month or even a year). Note that if cherry tip branch lengths are independently drawn from the same distribution, *d*_*i′*_ will be smaller than *d*_*i*_ for about half of the cherries. However, if some cherries are “notified” their real tip differences should be smaller than reshuffled ones. We therefore output the p-value calculated by the sign test.

As contacts upon notification might notify their own contacts, a scenario where a reshuffled cherry gets reconstructed from two real cherries belonging to the same contact tracing cluster is not impossible, especially when the values of *κ* (the number of contacts each index case might notify) and/or contact tracing probability *υ* are high. The branches of such a reshuffled cherry would not be independent and their sampling times could be closer in time, hence making detection of contact tracing in such cherries more difficult. This might slightly decrease our test sensitivity, but should not affect its specificity. In our experiments on simulated data the test worked well in all scenarios (see “CT test performance”). Real-life data might include other sources of non-random sampling than contact-tracing, for instance community-based testing campaigns [28]. As an index case and their potential contacts are likely to belong to the same community, we expect our cherry test to detect “contact tracing” in situations of community-based testing, which is certainly useful, but should not be misinterpreted.

## Mathematical formulation of BD and BD-CT(1) models

### Master equations and tree likelihood under BD model

The BD model can be described with master equations representing the likelihood densities of evolving as on the observed transmission tree, with time *t* going forward from the root (*t* = 0) till the time of the last sampled tip (*t* = *T*). These likelihood densities depend on the probability *U* (*t*) of an unobserved transmission tree that started from one individual at time *t* and evolved till *T*, without this individual nor any of their induced cases being sampled:

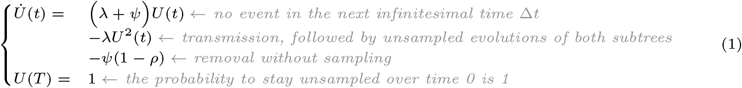

With the equation (1), we can write down the equation for the probability density *p*^(*i*)^(*t*) of evolving as on an observed tree branch *i* till time *t*_*i*_, starting on this branch at time *t ≤ t*_*i*_. The master equations for the BD model were initially developed by Stadler *et al*. [9], however what we present here is their branch-specific formulation that we proposed in [29]: In our formulation, *p*^(*i*)^(*t*) describes an evolution along a tree branch *i*, without taking into account the branch’s subtree. *t*_*i*_ corresponds to the time of the equation’s initial condition (typically at the end of the branch *i*).

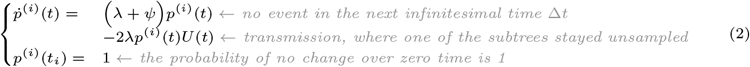

Using this equation, we can calculate the likelihood density *L*(𝒥|Θ) of a tree 𝒥under parameter values Θ = *{λ, ψ, ρ}* as:

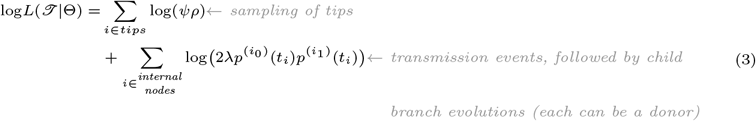

### Master equations under BD-CT(1) model

In the following we consider the simplest version of the BD-CT(*κ*) models family, where *κ* = 1 and hence only the last contact can be notified.

#### Tree branches under BD vs BD-CT(1) models

In the classical BD model with incomplete sampling, a branch of the tree corresponds to a virus evolution between the transmission corresponding to the node at the beginning of this branch and either a sampling event (if the branch is external) or a transmission corresponding to the node at the end of the branch (if it is internal). Note that since sampling is incomplete, hidden transmissions (leading to fully unsampled subtrees) could occur along this branch, where the unsampled subtree could correspond to either donor or recipient. Hence, we do not know whether the individual at the end of the branch (at time *t*_*i*_) is the same as at any time *t < t*_*i*_ along the branch. These two individuals could be different if a hidden transmission where the donor subtree stayed unobserved, occurred between times *t* and *t*_*i*_. *p*^(*i*)^(*t*) in equation (2) describes an evolution along such a tree branch, integrating over all the possibilities, starting from no hidden transmission along the branch and including any number of hidden transmissions with either donor or recipient tree staying unsampled (and hence potentially different individuals at times *t* and the branch end *t*_*i*_).

In trees generated by the BD-CT(1) model, however, the individuals do not behave in the same way when they are notified or not and, hence, other types of branches are possible. As in a real tree we do not know who might have notified whom, we have to integrate over all the possibilities to calculate the tree likelihood. We calculate the likelihood density of a tree using a pruning algorithm [30], while considering all the combinatorial possibilities for each node. For instance, for each tree tip we consider four possibilities: whether the corresponding individual was notified or not by someone, and on top of that, whether the tip’s individual notified or not their last contact upon sampling. Before describing the combinations behind tree likelihood calculation, we will introduce additional tree branch types.

#### Additional master equations under BD-CT(1) model

Let us consider a person A who got sampled at time *t*_*A*_ and successfully notified their contact B, who as a consequence got sampled at time *t*_*B*_ *≥ t*_*A*_. A and B hence correspond to tree tips, while their most recent common ancestor node AB corresponds to the transmission between A and B (see Fig. 2(1a)).

**Fig 2.**
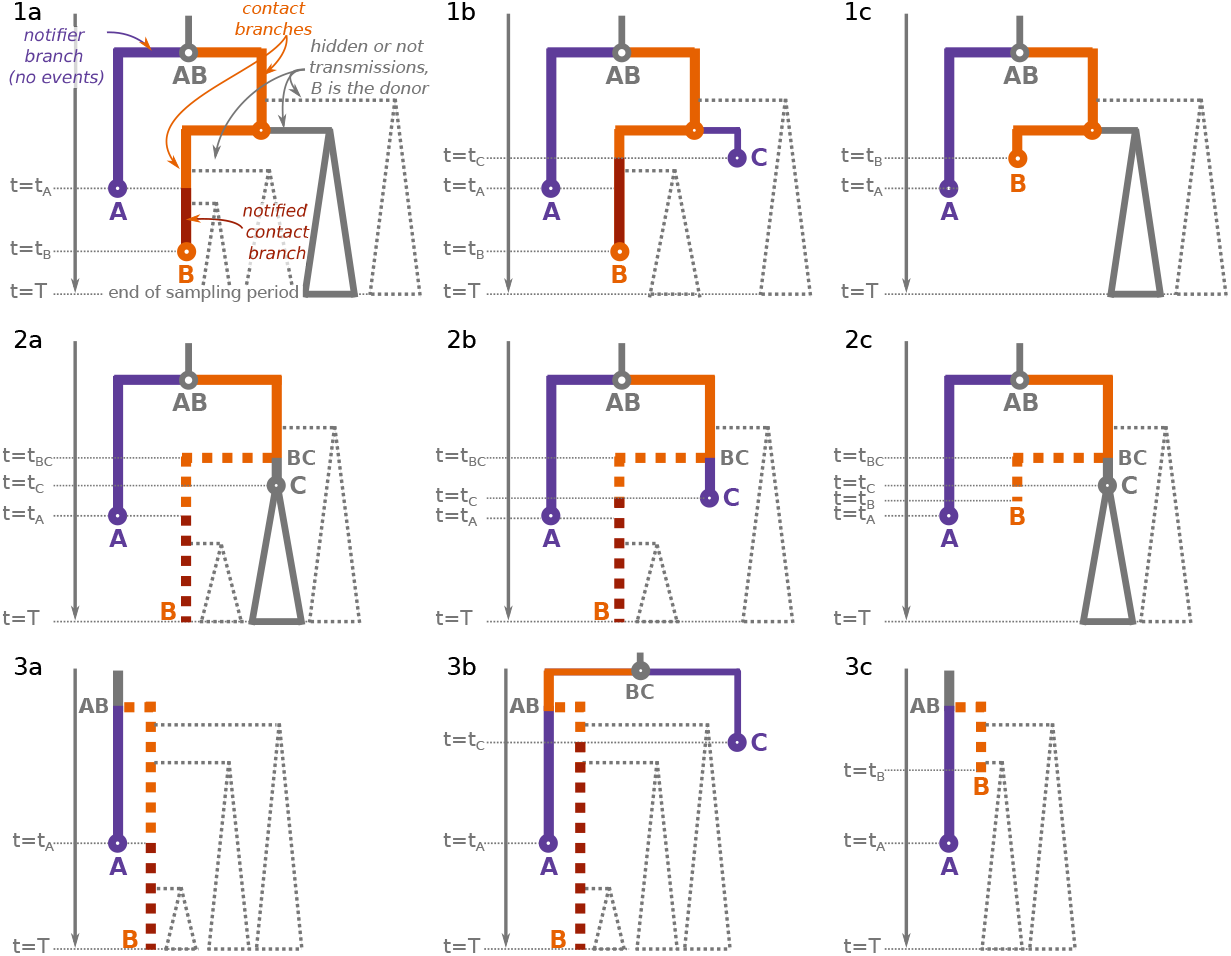
Examples of contact tracing in a transmission tree. We show 12 scenarios (one per panel) where an index individual A (violet) tried to notify their contact B (orange). The violet tip at time *t*_*A*_ represents the sampling of A, the internal node AB corresponds to the transmission between A and B. In row (**1**) B is observed by the end of the sampling period, in rows (**2, 3**) B is hidden. In row (**3**) also AB is hidden. In column (**a**) A was the first person to successfully notify B, in column (**b**) a third person C notified B before A did, in column (**c**) B was already removed via the standard procedure by the time A tried to notify them. The fact that A is the notifier of B implies that there is no hidden event along its branch AB-A (violet), and that in any transmission on the path AB-B (orange or dark-red) the donor is B. Moreover, between the moment of B’s first notification (*t*_*A*_ for **a**, *t*_*C*_ for **b**) and the moment of sampling of B (*t*_*B*_) the removal rate of B becomes *ϕ* instead of *ψ* (dark red). Finally, B might have been removed via a standard procedure before getting notified by their contact(s) (with rate *ψ* and a probability *ρ*, **c**). In the panels **3a-c** B’s subtree is fully hidden, and hence the transmission node AB is also unobserved. Therefore the A’s branch is composed of two parts, where the bottom part (violet) corresponds to A and contains no hidden transmissions, while the top part could correspond to A or not (e.g in **3b** it corresponds to B) and might contain hidden transmissions. In the panels **2a-b, 3a-b** B is hidden as by the end of the sampling period (*t* = *T*) B is not yet sampled, while in **2c, 3c** B is hidden as B got removed via the standard procedure without sampling before any notification. A’s branch is modeled with a notifier branch probability (4) in panels **1-2**, and with a mixed branch probability (see “S1 Appendix”) where the bottom part corresponds to the notifier (A) in panels **3**. If B did not notify their last contact upon sampling in panel **1**, branches on the path AB-B are modeled with a combination of contact probability before notification (orange, (5)) and contact probability after notification (dark red, (6)). Otherwise, the part after the transmission from B to their last contact is modeled with either a both notified and notifier branch probability (7) (as in **1a-b**) or a notifier branch probability (4) (as in **1c**), depending on the B’s notified status at that time. In the panels **2a-c**, the AB-C branch is modeled with a mixed branch probability (see “S1 Appendix”), where the top part corresponds to the contact state (B).

According to our model assumption 3, only the last contact can get notified. Hence, AB must be the parent node of A, and no hidden transmission is possible along the branch AB-A (as otherwise there would have been a contact of A who is more recent than B).

We will use a **notifier branch probability** 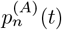 for such cases (when no hidden transmission occurred along the branch A, colored violet in Fig. 2):

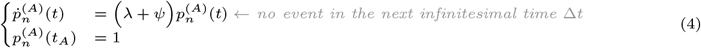

As B corresponds to A’s contact, the individual on the path between the transmission node AB and the tip B cannot change (is always B) and only transmissions from B to someone else are possible along the path leading from AB to B. These transmissions can be either hidden (where the recipient tree stayed unobserved) or observed: in both cases we know which subtree corresponds to the donor (the one that contains B). However, depending on whether B already got notified (*t ≥ t*_*A*_ in Fig. 2(1a)) or not yet (*t < t*_*A*_ in Fig. 2(1a)), B’s removal rate is different (is correspondingly *ϕ* or *ψ*). We will use a **contact branch probability before notification** 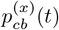 for cases when the individual at the end of the branch (or considered time interval, here *t*_*x*_ = *t*_*A*_) is the same as at time *t*, has not yet been notified, but might have transmitted to someone else (unobserved) along the way (colored orange in Fig. 2):

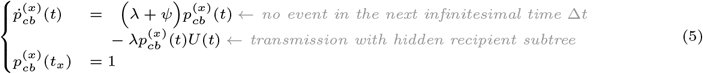

We will use a **contact branch probability after notification** 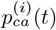 for cases when the individual at the end of the branch is the same as at time *t*, by time *t* has already been notified, and might have transmitted to someone else (unobserved) along the way (colored dark red in Fig. 2):

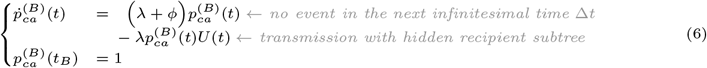

Finally, once sampled, B could have notified their own last contact (represented by the most recent hidden subtree in Fig. 2(1a)). This would imply that between the moment of transmission between B and their last contact (*t*_*x*_) and the moment of B’s sampling (*t*_*B*_) there was no event along the B’s branch. Hence, if B is a notifier, the part of the B’s branch after *t*_*x*_ should be modeled with a notifier probability instead (no event along the branch). Note that once B is notified, the removal rate becomes *ϕ*, and hence a **both notified and notifier branch probability** 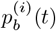 applies:

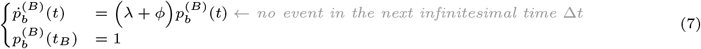

The equations 1, 2, 4-7 have the following closed form solutions:

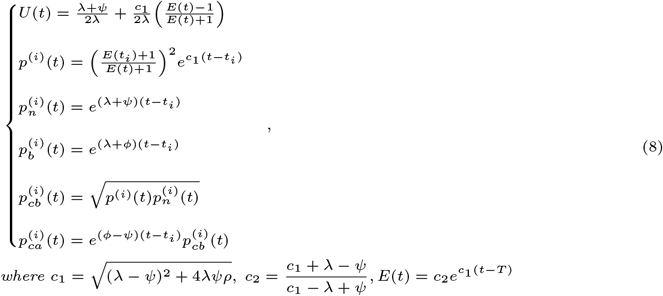

Note that on top of the scenario we just described (shown in Fig. 2(1a)), two other scenarios are possible for a person A who once sampled (at *t*_*A*_) notified their last contact B, who got sampled at time *t*_*B*_:

1b. by the time *t*_*A*_, B was already notified by someone else (e.g., at time *t*_*C*_ *< t*_*A*_ in Fig. 2(1b)).

This would lead to a shorter contact-before-notification part on the B’s branch (finishing at *t*_*C*_), and a longer notified-contact part (between *t*_*C*_ and *t*_*B*_).

1c. by the time *t*_*A*_, B was already sampled via a standard procedure (at time *t*_*B*_ *< t*_*A*_, hence the branch B would finish before the branch A, as in Fig. 2(1c)).

This would lead to the contact-before-notification part on the B branch finishing at *t*_*B*_ and no notified-contact part. Note that the branches between AB and B stay oriented as B remains the last contact of A.

Note that if A is a notifier, A is always observed, as notification only happens upon sampling. However, B could have stay unsampled: either because once notified (by A or another contact, who notified them before A did) B has not yet got sampled (Fig. 2(2a-b, 3a-b)); or because B got removed via the standard procedure without sampling before any notification (Fig. 2(2c, 3c)). Moreover, between the transmission AB and the moment of B’s notification/removal, B might have transmitted to someone else (e.g., C in Fig. 2(2), BC denoting the transmission). If that happened and someone in BC’s subtree got sampled, the branch AB-A would still correspond to the notifier A, while the other AB’s child branch would partially correspond to B (between AB and BC) and partially to C (starting at BC), as in Fig. 2(2). Otherwise (if there was no such transmission or all such subtrees stayed unobserved, as in Fig. 2(3)), the node AB will not be observed either, and A’s branch will get extended with its parent’s branch. Hence only the bottom part (after AB) of the A’s branch would correspond to the notifier probability (see Fig. 2(3)).

To model the cases when B is not sampled, we will need to describe the probability of a **mixed branch** 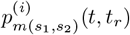 for branches that include a hidden contact subtree (AB-C branches in Fig. 2(2) and A’s tip branch in Fig. 2(3)). *s*_1_ here denotes the type of the top of the branch (e.g., between AB and BC in Fig. 2(2), contact) and *s*_2_ denotes the type of the bottom of the branch (e.g., between AB and A in Fig. 2(3), notifier). Let us denote the first notification time of the hidden contact as *t*_*r*_, and the time of the hidden contact subtree start as *t*_*h*_, while as usual *t*_*i*_ stands for the time at the branch end. For instance, in Fig. 2(2a) *t*_*i*_ = *t*_*C*_, *t*_*r*_ = *t*_*A*_, *t*_*h*_ = *t*_*BC*_; in Fig. 2(2b) *t*_*i*_ = *t*_*r*_ = *t*_*C*_, *t*_*h*_ = *t*_*BC*_; in Fig. 2(3c) *t*_*i*_ = *t*_*r*_ = *t*_*A*_, *t*_*h*_ = *t*_*AB*_.

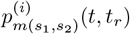 combines three elements: the probability of the top part of the branch 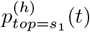, which finishes with a transmission to or from the hidden contact, the probability of the bottom part of the branch 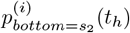 (without taking into account the event at its end) and the probability of the hidden contact subtree *U*_*p*_(*t*_*h*_, *t*_*r*_). We describe the details behind calculation of 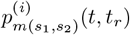 in S2 Appendix.

In real trees we do not know sampled individual’s status (individual who did not notify, notifier, contact notified in time or not), and hence during the tree likelihood calculation, we will have to consider all these scenarios.

#### Tree likelihood under BD-CT(1) model

Using the branch probability formulas described above, we can calculate the likelihood density of a tree using a pruning algorithm [30]. For each visited node *i* we calculate the likelihood density of its subtree including the branch *ji* connecting *i* to its parent node *j* for two cases depending on *i*’s receiving-a-notification status (shown with the first superscript, 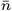 or ^*n*^). In the first case, 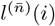, the node *i* corresponds to an unnotified individual. In the second case, *l*^(*n*)^(*i, r*), the branch *ji* and the node *i* correspond to an (eventually) notified contact (notified by the individual corresponding to a tip *r* at time *t*_*r*_). If *i* is a tip, we additionally consider whether there is an observed contact in our tree, whom the individual *i* notified (shown with the second superscript, ^*n*^ or 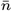). For instance, 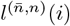 represents the case where the tip *i* was not notified but notified their last contact (who is observed).

The tree 𝒥 likelihood density *L*(𝒥| Θ) under BD-CT model with parameters Θ = *{λ, ψ, ρ, ϕ, υ}* can be calculated as 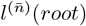, as the root can only correspond to an unnotified individual (since there was no observed transmission before it).

We describe the subtree likelihood density calculation case by case in S3 Appendix.

#### Extension to forests

As we previously described in [29], the likelihood calculation with MTBD models can be easily extended to forests. This extension applies also to the BD-CT models. Forests are useful for cases when a (sub-)epidemic started with several infected individuals (e.g., due to multiple pathogen introductions to a country of interest or due to a change of health policies leading to a change in parameter values). In this case the (sub-)epidemic leads to a forest ℱ of *f* observed trees: 𝒥_1_, …, 𝒥_*f*_. The forest ℱ might also include a certain number *u* of unobserved trees, for which none of their tips got sampled. Forest likelihood formula hence combines the likelihoods of *f* observed and *u* hidden trees, and can be represented in a logarithmic form:

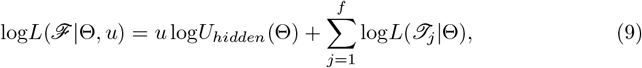

where *U*_*hidden*_(Θ) = *U* (*t*_*start*_) and *t*_*start*_ is the (potentially averaged) start time of the hidden trees.

For given model parameter values Θ we can estimate the number of hidden trees *u* from the number of observed trees *f* as:

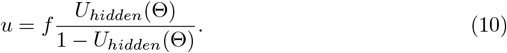

The tree likelihood formula is a special case of equation (9), where *f* = 1 and *u* = 0.

### BD and BD-CT(1) parameter and CI estimators

We implemented a maximum-likelihood parameter estimator for the BD-CT(1) model (which we called bdct). It estimates the BD-CT(1) model parameters Θ = (*λ, ψ, ϕ, ρ, υ*) ∈ ℝ^5^ for a forest ℱ comprising *f ≥* 1 observed trees, provided one of the BD parameters (*λ, ψ* or *ρ*) is fixed (for identifiability reasons). We also implemented the classical BD model estimator, bd.

Once the optimal parameter values are found, we calculate their confidence intervals (CIs) using Wilks’ method [31]. For each non-fixed parameter *p ∈* Θ, we calculate its 95%-CI as 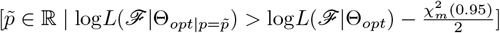 where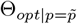 is the maximum-likelihood value for the other non-fixed parameters when 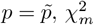 (0.95) is the value of chi-squared distribution with *m* degrees of freedom corresponding to the significance level of 0.95, and *m* is the number of free model parameters. For BD-CT(1) with *ρ* being fixed, *m* = 4 and 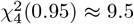. For BD with *ρ* being fixed, *m* = 2 and 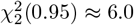.

## Code availability

Our parameter estimators are implemented in Python 3. They use ETE 3 framework for tree manipulation [32] and NumPy package for array operations [33].

They are available as command-line programs (*bd infer, bdct infer*) and a Python 3 package via PyPi ([bdct](https://pypi.org/project/bdct)), and via Docker/Singularity (evolbioinfo/bdct). The source code, installation and usage documentation are available on GitHub at github.com/evolbioinfo/bdct.

The parameter estimator package also includes the cherry test (*ct test*, see section “Testing for presence of Contact Tracing”).

### Zurich men-having-sex-with-men HIV-1 B tree and sampling probability

We applied our estimators to asses the contact tracing for HIV-1 subtype B epidemic among men-having-sex-with-men (MSM) in Zurich. We used the phylogenetic tree of 200 samples reconstructed by Rasmussen *et al*. [20] from the sequences collected as a part of the Swiss Cohort Study [34] between 1988 and 2014. This tree was also previously analyzed by Voznica *et al*. [21] using a Birth-Death model with SuperSpreading (BDSS). Following Voznica *et al*. [21], we fixed the sampling probability for this tree at 25%: *ρ* = 0.25.

### UK HIV-1 B forest reconstruction and sampling probability estimation

To asses the contact tracing in HIV-infected patients in the UK, we used the phylogenetic tree from our recent study of HIV drug resistance in the UK [35]. The tree tips represent viruses sampled from 40 055 individuals between 1996 and 2016. The samples used for the tree reconstruction were obtained from the UK HIV Drug resistance database [36]. The sampling date metadata for these samples were expressed in months (e.g., Aug 2015), hence potentially creating simultaneous sampling dates for cases that were sampled on different days of the month. To avoid this issue we randomly selected a date within the specified month for each sample, and time-scaled the tree with LSD2 [37] (v2.4.1, strict molecular clock with outlier removal). We repeated random date resolution and time-scaling 10 times, obtaining 10 trees with slightly different branch lengths and root dates between Oct 5 and Nov 11, 1962.

Between 2012 and 2015, in the UK antiretroviral treatment (ART) of HIV-infected individuals was initiated when their CD4 count dropped below 350 cells/mL [38]. However “if a patient with a CD4 cell count *>* 350 cells/mL wishes to start ART to reduce the risk of transmission to contacts, this decision is respected and ART is started” [38]. Before 2012, treatment started with an even lower CD4 (i.e., later). The British HIV Association 2015 guidelines recommend all individuals with suspected or diagnosed primary HIV infection “are offered immediate ART” [39]. To have a more homogeneous setting in terms of access to ART, awareness of the HIV infection, etc., we cut the dated trees between 2012 and 2015.

To estimate the BD-CT parameter values, we needed to fix one of the parameters for model identifiability. We therefore estimated the base sampling probability *ρ* as follows. According to the [“Towards elimination of HIV transmission, AIDS and HIV-related deaths in the UK”](https://webarchive.nationalarchives.gov.uk/ukgwa/20181112132123mp_/https://assets.publishing.service.gov.uk/government/uploads/system/uploads/attachment_data/file/602942/HIV_in_the_UK_report.pdf) report, the estimated total number of people living with HIV in the UK in 2015 was 101 200. According to our estimate [35], 66.5% of the HIV-infected individuals in the UK were infected with HIV-B. We hence estimated that in 201567 298 people were living with HIV-B. Our trees contained 39 047 tips sampled by the end of 2015, and therefore account for about 58% (= 39 047*/*67 298 · 100%) of the total number of individuals living with HIV-B in the UK. We hence fixed the sampling probability *ρ* to 0.58.

## Results

### CT leaves clear traces in transmission trees

#### CT test performance

To illustrate the CT test performance, we applied it to the twelve 100-tree simulated datasets, corresponding to the three classical MTBD-family models: BD, BDEI (Birth-Death Exposed-Infectious [13]) and BDSS (Birth-Death with Super-Spreading [12]), and to their −CT versions with *κ* = 1 (possibility to notify the latest contact), *κ* = 2 (possibility to notify the last two contacts), and *κ* =*∞* (possibility to notify all the contacts), see “MTBD-CT tree simulator and test datasets” for dataset details. The results are summarized in Table 1.

**Table 1.**
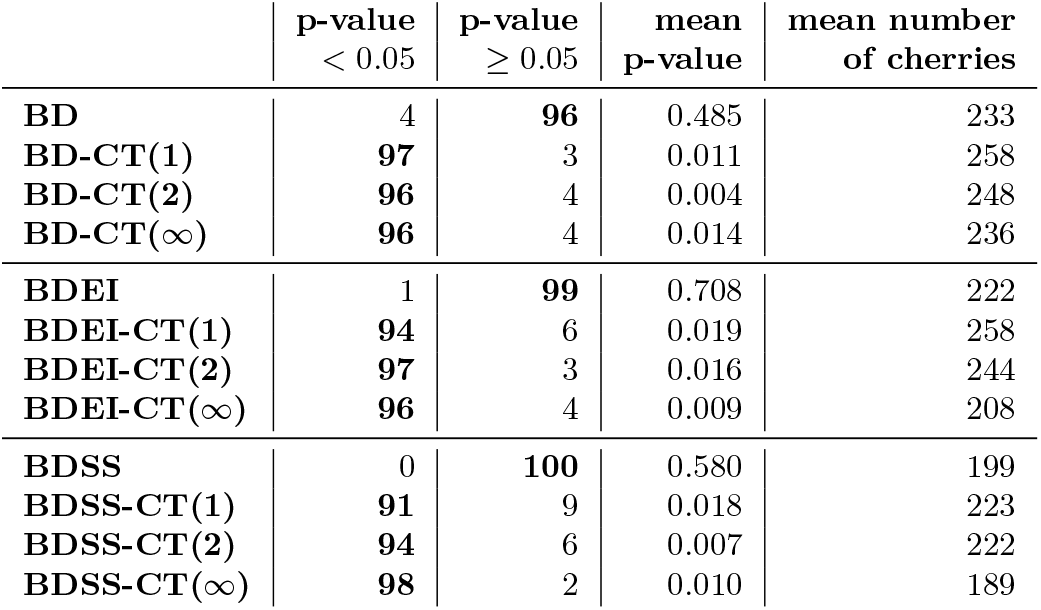
CT test performance on trees generated under BD, BDEI, BDSS models and their −CT versions (100 trees per model, each having 500-1000 tips). We report the number of trees for which the CT-test p-value was *<* 0.05, *≥* 0.05, the mean p-value and the mean number of cherries for each dataset.

For 96/99/100 out of 100 trees in the BD/BDEI/BDSS dataset the CT test p-value was above 0.05 (i.e., 96%/99%/100% true negative and 4%/1%/0% false positive results).

For 97/94/91 out of 100 trees in the BD-CT(1)/BDEI-CT(1)/BDSS-CT(1) dataset the CT test p-value was below 0.05 (i.e., 97%/94%/91% true positive and 3%/6%/9% false negative results). It showed similar performance for higher values of *κ*.

Our test overall showed high specificity (98% on the combined 300-tree dataset without CT) and good sensitivity (95% on the combined 900-tree CT dataset).

#### Likelihood ratio test (LRT) performance

For the BD and BD-CT datasets we performed an LRT with the parameters estimated by our bd and bdct estimators: *−*2(*l*_*BD*_*− l*_*BDCT*_) ≶ 5.99 (where 5.99 is the value of the chi-squared distribution with 2 degrees of freedom corresponding to a significance level of 0.95). The test selected the correct model (-CT vs no CT) for 97%, 99%, 100% and 99% of the corresponding BD, BD-CT(1), BD-CT(2) and BD-CT(*∞*) trees.

These results and those obtained with the CT test show that the trees generated with the CT component are very different from those generated without it. It should also be noted that likelihood calculation and LRT performed using bdct seem to be robust with respect to the value of *κ* (good results are obtained when *κ* = 2 and *κ* = *∞*).

### Accounting for contact tracing is crucial for accurate parameter estimation

We assessed the performance of our maximum-likelihood estimators for the BD model (bd) and BD-CT(1) model (bdct) on four simulated datasets, each containing 100 trees (see “MTBD-CT tree simulator and test datasets” for details), simulated under either the classical BD model or its −CT(1), −CT(2) or −CT(*∞*) versions. We applied our parameter estimators to each of the trees fixing the sampling probability *ρ* to its true value for identifiability.

As expected, the tree likelihoods for the parameter values estimated by bd on the BD trees, and by bdct on the trees from all the datasets, were higher than or equal to the tree likelihoods for the true parameter values for these trees. This shows the consistent behavior of our likelihood calculation and optimization.

#### Point estimates

The estimation results are shown in Fig. 3. We report average relative errors and biases 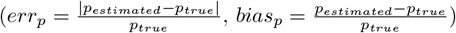 for the rate parameters (*p ∈ {λ, ψ, ϕ}*) and average absolute errors and biases (*err*_*υ*_ = |*υ*_*estimated*_ *−υ*_*true*_|, *bias*_*υ*_ = *υ*_*estimated*_ *−υ*_*true*_) for the contact tracing probability *υ*, which can be very small.

**Fig 3.**
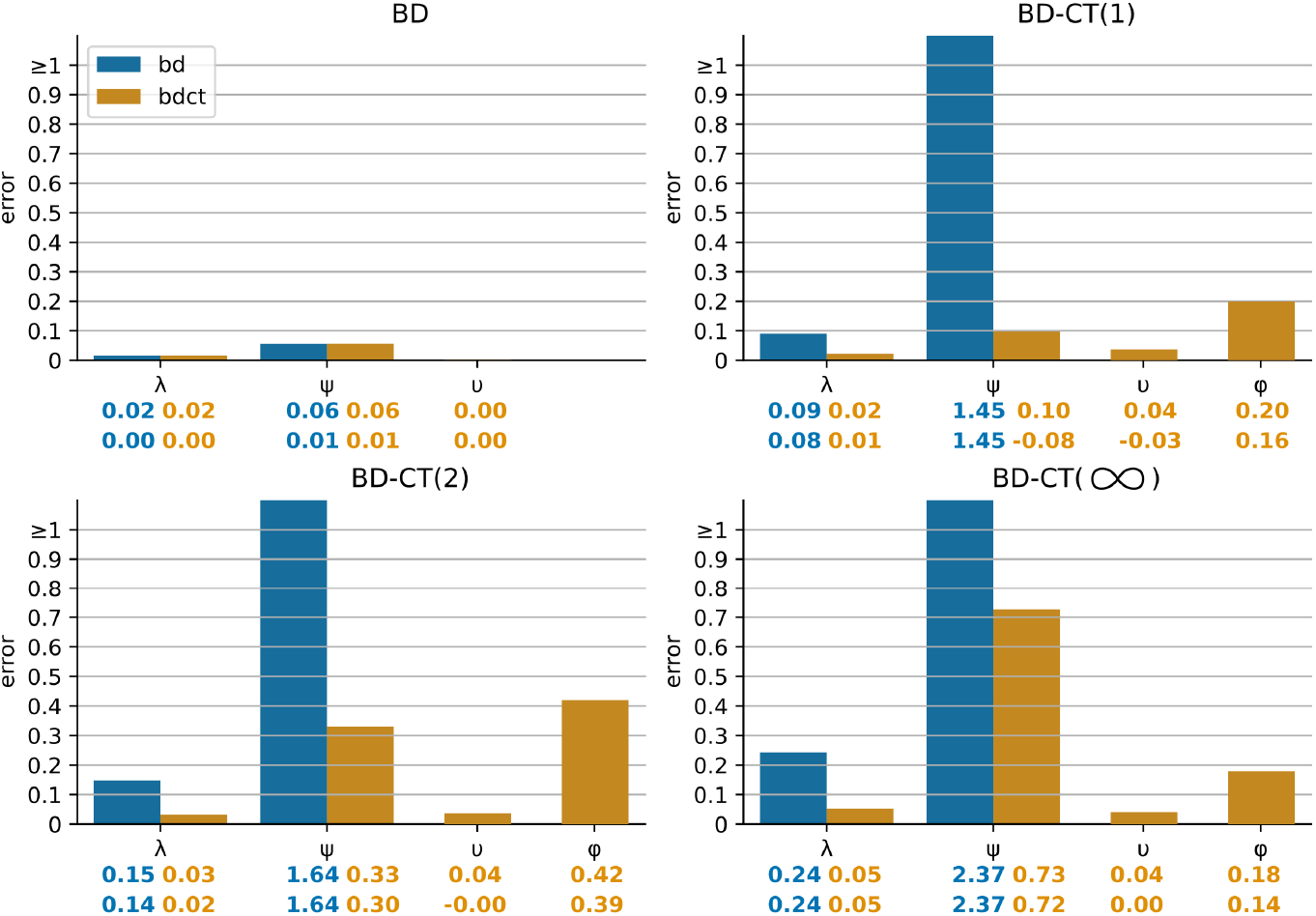
Comparison of inference accuracy of bd (blue) and bdct (yellow) estimators on datasets of 100 trees of 500–1 000 tips generated under the BD (top left), BD-CT(1) (top right), BD-CT(2) (bottom left), and BD-CT(*∞*) (bottom right) models, with *ρ* fixed to its true value. We show the bar-plots (colored by estimator) representing mean errors for each parameter. The error values are displayed below the corresponding bar-plots. Below them the mean bias values are displayed. For the rate parameters relative errors and biases are shown (distance for errors, difference for biases, between the estimate and the true value divided by the true value), for the contact tracing probability *υ* absolute errors and biases are shown (as for the BD model the true value is zero). For *ϕ* only the cases with non-zero estimates of *υ* were considered.

The estimators performed well on the models that correspond to them (BD for bd, BD or BD-CT(1) for bdct): the average relative errors for the BD-model parameters are within 10%, the average absolute error for *υ* (with prior within [0.01, 0.75]) was less than 0.04. However, the performance of our estimators got worse when the model was misspecified (presence of CT for bd, *κ >* 1 for bdct).

Interestingly, contact tracing had different effects on the estimation of different parameters. Not accounting for contact tracing at all led to a drastic overestimation of the standard removal rate *ψ* (e.g., relative bias of 145% and 237% for bd on BD-CT(1) and BD-CT(*∞*) trees, respectively). When accounting for less contacts than in the true model, the bias is less drastic but still significant (e.g., relative biases of 30% and 72%, for bdct on BD-CT(2) and BD-CT(*∞*) trees). Not accounting for contact tracing also led to a slight overestimation of the transmission rate *λ* (e.g., relative bias of 24% for bd and of 5% for bdct on BD-CT(*∞*) trees). Intuitively, in the absence (or decreased presence in case of wrong *κ*) of a contact tracing mechanism, which allows for faster removal and sampling of a fraction of the infected population, the compensation mechanism is to increase the standard removal rate for the entire population. Both effects were much less present for the bdct estimator than for bd, i.e., when one contact was allowed to be traced. Combined with the performance of bdct (equivalent to bd’s) on the BD dataset, this advocates for using bdct in all the situations to at least reduce the estimation bias. Interestingly, the estimation of *υ* remained accurate (average absolute error *<* 0.04) and unaffected by model misspecification.

The notified sampling rate *ϕ* was the hardest parameter to estimate in all the CT settings (average relative error of 18-42%, and a bias towards overestimation in all the settings). The difficulty in its estimation is not surprising as there is the least information on it in the trees. For instance, the contact tracing probability *υ* comes into play (positively or negatively) on every sampling event (i.e., every tree tip), while the notified sampling rate *ϕ* only participates in those of these cases where the index (i) decides to notify (i.e., the positive draw of *υ*) and (ii) gets removed before their notified contact. Moreover, the bias in the estimation of this parameter could be due to the approximations in the BD-CT(1) likelihood calculation that we had to make in order to obtain a closed-form solution (e.g., setting hidden contact notification times to the middle of the corresponding intervals, see S2 Appendix).

#### Confidence Intervals (CIs)

The percentage of the trees for which true parameter values *p*_*true*_ were within the CI estimates and relative CI widths 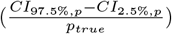 are shown in Table 2. For the CIs as for the point estimates, bdct and bd estimators performed equally well on the BD trees (100% of estimates within the CIs), however the CIs estimated by bdct were slightly wider (15% and 49% for respectively *λ* and *ψ* versus 12% and 39% for bd). On the BD-CT(1) trees the bd estimator performed significantly worse (only 51% and 7% of CIs included the true values of respectively *λ* and *ψ* versus 93% and 89% for bdct). As *κ* increased, the performance of bd dropped (down to 22% for *λ* and 7% for *ψ* on BD-CT(*∞*) trees). The performance of bdct with respect to *λ* and *ψ* also decreased with the increase of *κ*, but less drastically than for bd (reaching 84% and 36%, respectively, on BD-CT(*∞*) trees). This finding is somewhat expected due to the bias in the point estimates for *λ* and especially *ψ*. Interestingly, CI performance for *ϕ* and *υ* remained stable or even improved with the increase of *κ* (95% and 91%, respectively, on BD-CT(*∞*) trees). As with the point estimates, *ψ* was most affected by contact-tracing model misspecification, and much more so when contact tracing was not included at all (bd) than when it was partially included (bdct, *κ* = 1).

**Table 2.** Percentage of simulated BD, BD-CT(1), BD-CT(2) and BD-CT(*∞*) trees for which the true parameter values were within the bd and bdct-estimated 95% CIs, and the median CI width: 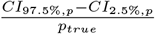. (The sampling probability *ρ* was fixed to its true value in all the settings.)

| true model | parameter | bd estimator |  | bdct estimator |  |
| --- | --- | --- | --- | --- | --- |
|  |  | % within CI | median CI width | % within CI | median CI width |
| BD | $\lambda$ | 100% | 11.5% | 100% | 14.6% |
| | $\psi$ | 100% | 39.0% | 100% | 49.1% |
| | $v(=0)$ | — | — | 100% | 0.03* |
| BD-CT(1) | $\lambda$ | 51% | 12.6% | 100% | 15.2% |
| | $\psi$ | 7% | 66.6% | 98% | 55.5% |
| | $\phi$ | — | — | 93% | 49.6% |
| | $v$ | — | — | 90% | 35.5% |
| BD-CT(2) | $\lambda$ | 38% | 14.5% | 97% | 16.7% |
| | $\psi$ | 11% | 59.2% | 63% | 56.9% |
| | $\phi$ | — | — | 92% | 54.4% |
| | $v$ | — | — | 90% | 38.4% |
| BD-CT(∞) | $\lambda$ | 22% | 15.9% | 84% | 17.6% |
| | $\psi$ | 7% | 68.4% | 36% | 67.4% |
| | $\phi$ | — | — | 95% | 55.8% |
| | $v$ | — | — | 91% | 40.6% |

\*absolute width ( $CI_{97.5\%,v} - CI_{2.5\%,v}$ ) is given instead of the relative one, as the true value is zero.

## Application 1: Men-having-sex-with-men HIV-1 B epidemic in Zurich

We applied our estimators to assess the presence of contact tracing for HIV-1 subtype B epidemic among men-having-sex-with-men (MSM) in Zurich, using a 200-sample time-scaled phylogenetic tree from the study by Rasmussen *et al*. [20], and fixing the sampling probability to 25%: *ρ* = 0.25 (see “Zurich men-having-sex-with-men HIV-1 B tree and sampling probability” for more details).

The CT test detected contact tracing in this tree (p-value of 0.014). The likelihood ratio test also suggested a BD-CT model: log-likelihood under the bdct-estimated parameters was *l*_*bdct*_ = *−*1 140 vs. *l*_*bd*_ = *−*1 146 under the bd-estimated parameters, which gives an LRT statistic of 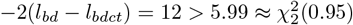. Our bdct parameter estimator inferred a contact tracing probability *υ* = 0.09 CI [0.001 *−* 0.314], *R*_*e*_ = 1.43 [1.17 *−* 1.76], infectious time 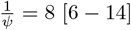 years, and 4.9 months [3 days – 2.3 years] between contact notification and their sampling. These results globally agree (CIs intersect) with the *R*_*e*_ estimate for this data using a coalescent model from Rasmussen *et al*. [20] (between 1.0 and 2.5), and the estimates using the BDSS model from Voznica *et al*. [21]: *R*_*e*_ of 1.6 and 1.7 and an infectious period of 10.2 and 9.8 years (depending on the tree representation used for the deep learner).

Note that this tree covers a long non-homogeneous time interval (between 1974, the root of the tree, and 2012), including times with no or limited access to treatment, when the infectious time corresponded to progression to AIDS (*≈* 10 years), as well as later times, when antiretroviral treatment (ART) became available, and hence the infectious period would correspond to the time till ART start, as, when taken properly, it prevents transmission. Our estimate (8 years) represents a mix of these scenarios. This also holds for the other estimated parameters, whose values represent a somewhat averaged estimate over the time period. As this dataset was already small, we did not cut it to obtain a more homogeneous treatment policy (as we did in Application 2).

Moreover, as our results on the simulated data suggest, not accounting for (enough) contact tracing (i.e., estimating BD-CT(1) parameters on the data generated under*κ >* 1) leads to overestimation of *ψ* and hence underestimation of the infectious time 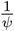 and potentially 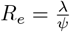. Our estimate should therefore be treated as a lower bound (i.e., an infectious time of at least 8 years and a reproduction number of at least 1.43). In the same vein, parameter inference with the bd estimator gave a slightly lower estimate of *R*_*e*_ and a shorter infectious time, though with intersecting CIs (*R*_*e*_ = 1.32 [1.16 *−* 1.51], 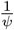= 7 [6 *−* 9] years).

We could not find estimates of the percentage of HIV infections detected through contact tracing in Switzerland with which to compare our estimation of *υ*. An editorial in “Revue médicale Suisse” [40] stated in 2004 that there was no systematic assisted contact tracing for HIV in Switzerland. This situation might have improved by 2012. In addition, there might have been patient referral (patients notifying their contacts on their own). Overall, this supports a relatively low value for *υ*.

## Application 2: HIV-1 B epidemic in the UK

We also applied our estimator to asses the presence of contact tracing in HIV-1 subtype B infected individuals in the UK between 2012 and 2015. This time period was chosen to represent a homogeneous treatment policy (treatment start at CD4 count below 350 cells/mL [38]). We used 10 forests reconstructed from HIV-1 B sequences from the UK HIV Drug resistance database [36] (see “UK HIV-1 B forest reconstruction and sampling probability estimation” for details).

The CT-test p-values were below 0.05 for all the forests (between 9 · 10^*−*20^ and 1 · 10^*−*14^), strongly suggesting the presence of contact tracing. The likelihood ratio test also suggested a BD-CT model: For instance, on forest 1, log-likelihood under the bdct-estimated parameters was *l*_*bdct*_ = − 28 984 vs. *l*_*bd*_ = *−* 29 002 under the bd-estimated parameters, which gives the LRT statistic of 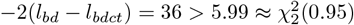.

Using sampling probability *ρ* = 0.58 (see “UK HIV-1 B forest reconstruction and sampling probability estimation” for its estimation), we estimated the contact tracing probability *υ* = 0.01 CI [0.002 *−* 0.015], *R*_*e*_ = 1.2 [1.20 *−* 1.29], infectious time 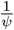 = 2.3 [2.24 − 2.42] years, and 8 [2 − 17] days between contact tracing and their sampling. The results were consistent between 10 forests and are shown in Table 3.

**Table 3.** BD-CT model parameters estimated for UK HIV-1 B epidemic between 2012 and 2015, assuming that our data represents 58% of infected cases (*ρ* = 0.58).

| fo-<br>rest | num.<br>tips | num.<br>trees | $R_e$ | infectious<br>time<br>$\frac{1}{\psi}$ [years] | contact<br>removal t.<br>$\frac{1}{\phi}$ [days] | $\nu$ |
| --- | --- | --- | --- | --- | --- | --- |
| 1 | 11 117 | 8 838 | 1.24 [1.20 – 1.29] | 2.34 [2.25 – 2.42] | 8 [4 – 15] | 0.008 [0.004 – 0.015] |
| 2 | 11 109 | 8 833 | 1.24 [1.20 – 1.29] | 2.33 [2.25 – 2.42] | 7 [2 – 14] | 0.007 [0.003 – 0.013] |
| 3 | 11 122 | 8 844 | 1.24 [1.20 – 1.29] | 2.34 [2.25 – 2.42] | 8 [4 – 13] | 0.008 [0.004 – 0.014] |
| 4 | 11 119 | 8 838 | 1.24 [1.20 – 1.29] | 2.34 [2.25 – 2.42] | 8 [4 – 14] | 0.008 [0.004 – 0.015] |
| 5 | 11 116 | 8 829 | 1.24 [1.20 – 1.29] | 2.34 [2.25 – 2.42] | 9 [4 – 17] | 0.007 [0.002 – 0.014] |
| 6 | 11 111 | 8 827 | 1.24 [1.20 – 1.29] | 2.34 [2.25 – 2.42] | 8 [4 – 15] | 0.007 [0.002 – 0.013] |
| 7 | 11 119 | 8 838 | 1.24 [1.20 – 1.29] | 2.33 [2.24 – 2.42] | 8 [4 – 15] | 0.007 [0.002 – 0.013] |
| 8 | 11 111 | 8 830 | 1.24 [1.20 – 1.29] | 2.33 [2.24 – 2.42] | 7 [3 – 14] | 0.007 [0.003 – 0.013] |
| 9 | 11 120 | 8 833 | 1.24 [1.20 – 1.29] | 2.33 [2.24 – 2.42] | 8 [3 – 15] | 0.007 [0.002 – 0.013] |
| 10 | 11 112 | 8 827 | 1.25 [1.20 – 1.29] | 2.34 [2.25 – 2.42] | 8 [3 – 14] | 0.008 [0.003 – 0.013] |

In the UK in 2015 the majority of infected population (72% [41]) was on antiretroviral treatment, which, when taken properly, prevents transmission. Hence, the average infectious time roughly represents the average time between infection and treatment start. Note that, according to our results on simulated data, this time might be underestimated if the true *κ* value (number of partners that can be notified) is greater than one. Hence, our result suggests that the average time between infection and treatment start is *at least* 2.3 years.

According to the HIV report from the Public Health England [19], in England in

2013, less than 3% of sampled infected individuals were detected through contact tracing. Our model considers only contacts that caused a virus transmission, while in real life partner notification may permit detection of seropositive individuals with whom there was no transmission. This explains why our estimate is even lower (1%). It should be interpreted as 1% of sampled infected individuals were detected through contact tracing between the transmitting and the acquiring partner.

For comparison, parameter estimation on the same forests with the BD model gave similar results (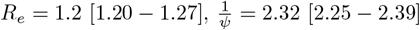 years), however the infectious duration was slightly shorter, which is in line with overestimation of the removal rate when not accounting for contact tracing (which we have seen on simulated data).

As expected the estimates on the much smaller Zurich dataset show larger CIs than those on the larger UK one. The CIs estimated on the two datasets intersect for the effective reproduction number *R*_*e*_, while the infectious time is larger for Zurich (CI of 6 *−* 14 years vs 2.2 *−* 2.4 years for the UK). This is however expected as the Zurich tree covers a longer, earlier and non-homogeneous time interval (between 1974 and 2012), including times with no or limited access to treatment, when the infectious time corresponded to progression to AIDS (*≈* 10 years). The CIs for contact tracing probability *υ* and contact sampling time 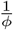 also intersect on the two datasets, though they do not have to, as the contact tracing policy might differ between the two countries, especially during the long and non-homogeneous time interval represented by the Zurich tree. The point estimates suggest stronger contact tracing probability and longer notified contact sampling time in Zurich than in the UK. Longer delays in contact testing might be explained by the absence of assisted partner notification in Switzerland.

## Discussion

We proposed an extension of the phylodynamic birth-death model that accounts for non-random sampling due to contact tracing (BD-CT). In real-life epidemics, detected infected individuals might notify the people whom they think they might have infected or got infected by, and who in turn might get tested and detected much faster than via the standard procedure. Our model accounts for this scenario.

The −CT(*κ*) extension adds two constant parameters to the classical BD model: the probability of contact tracing *υ* and the contact detection rate *ϕ*. The meta-parameter *κ* controls for the number of last contacts that may be traced by each index case. The −CT(*κ*) extension can also be applied to a more general, multi-type birth-death (MTBD) modeling framework, which accounts for heterogeneity in the infected population.

We developed a statistical CT-test allowing to distinguish between trees generated under models with and without contact tracing. Its application to simulated data showed high sensitivity and specificity.

We developed a mathematical framework for calculation of the likelihood density of a transmission tree (approximated by a time-scaled phylogenetic tree) under the BD-CT(1) model, and implemented a maximum-likelihood parameter estimator, bdct, based on this framework. For comparison we also implemented a classical BD model estimator, bd. The advantage of the BD-CT(1) model is that the differential equations describing it have closed-form solutions, which are quick to calculate and do not require numerical approximation. This is not the case for more complex MTBD(-CT) models. Moreover we did not find a closed-form solution even for the BD-CT(*κ*) models with *κ >* 1. Different *κ* values should be considered in future work.

Our tests on simulated data showed that our estimators robustly infer model parameter values and their CIs in the setting when the base sampling probability *ρ* is fixed and the model is correct (i.e., BD-CT(1) for bdct, BD for bd). Fixing one of the model parameters is needed for model identifiability and, in practice, *ρ* may often be approximated from epidemiological data, e.g., as the proportion of sampled cases among the declared ones.

Importantly, our simulations show that in the presence of tracing of the last contact (i.e., the BD-CT(1) model), the classical BD model estimator bd fails to accurately infer epidemiological parameters. On the contrary, in the absence of contact tracing (the BD model), the bdct estimator correctly identifies the parameter values, and estimates a very low contact tracing probability *υ*, whose confidence intervals include zero. The performance of both bd and bdct gets worse with the increase of *κ* (possibility to notify more than one last contact). In particular, not (fully) accounting for contact tracing leads to overestimation of the standard removal rate *ψ* (and hence underestimation of the infectious duration 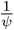). Interestingly, the estimation of the contact tracing probability *υ* seems to be robust against *κ* misspecification. While keeping in mind overestimation of *ψ* under *κ* misspecification, it is still useful to account for contact tracing at least partially: For instance, in the setting when the last two contacts can be notified (i.e., the BD-CT(2) model), parameter CIs inferred by bdct (i.e., the BD-CT(1) model) contain the true values of the transmission rate *λ* and the removal rate *ψ* in respectively 97% and 63% of cases, while not accounting for contact tracing at all using the bd estimator leads to the CIs containing them only in 38% and 11% of cases.

We applied the CT-test and our estimator to the HIV-1 B MSM cluster in Zurich and to the HIV-1 B epidemic in the UK between 2012 and 2015. The test detected presence of contact tracing in both cases. The obtained estimates of epidemiological parameters are in agreement with what we now know about the HIV-1 B epidemic (e.g., *R*_*e*_ between 1.2 and 1.8). We estimated a modest contact tracing probability (9% in Zurich, 1% in the UK), and rapid notified contact detection (*≈* 5 months vs *≈* 8 years for detection under the standard procedure in Zurich; and *≈* 8 days vs *≈* 2.3 years for detection under the standard procedure in the UK). Note that the contact tracing probability represents “successful notification”, which combines several conditions: the index patient remembers and agrees to notify the contact, the contact is reached, agrees to take a test, is indeed infected, and moreover the transmission happened during the interaction between the index and the contact. This explains the seemingly low values. We applied our BD-CT(1) estimator to these data, however as our simulations show, allowing for tracing of only the last contact might lead to underestimation of the infectious time, so our estimates of 8 years for Zurich and 2.3 years for the UK should be treated as lower bounds. Moreover, other aspects of these epidemics (e.g., potential super-spreading events as in BDSS-CT models, or change of health policies and hence parameter values over time, as in MTBD-Skyline models [15]), which we did not account for in this study, might also impact the parameter estimation, and should be incorporated in the future work.

While we only applied our parameter estimator to sexually transmitted infections (and in particular HIV), contact tracing is not limited to them. For instance it was successfully used to reduce the spread of SARS-CoV-2 [42] and Ebola [43]. Combining genomic sequence data with MTBD-CT models could give additional insights on impact of contact tracing on slowing these epidemics. However, as these viruses feature an incubation period, they would require BDEI-CT or even BDEISS-CT (combining incubation and super-spreading) estimators instead of the BD-CT one. The BD-CT(*κ*) model is the first step towards accounting for non-random sampling in phylodynamic models. In order to reduce its mathematical complexity (having closed-form solutions for its differential equations) and make parameter estimation fast, we combined several conditions of the complex contact tracing process into one “successful notification” probability *υ* (see above), and made several additional assumptions: (i) only sampled individuals can notify their contacts; (ii) only last *κ* contacts can get notified; (iii) once notified, contacts are always sampled upon removal (however, potentially after the end of the considered sampling period). These assumptions may be too simple to fully grasp the real contact tracing process and should be relaxed in the future work. In particular, as our simulations show, only accounting for *κ* = 1 as we did in this study can lead to overestimation of the standard removal rate. Another needed extension should account for population heterogeneity (i.e., MTBD-CT), which would incorporate such aspects of epidemics as incubation and super-spreading. Finally, it would also be useful to incorporate the possibility of parameter change over time as the health policies get updated, including for the parameters related to contact tracing. This could be modeled by extending MTBD-Skyline models [15] with contact tracing. As our experiments show, it is much easier to implement a tree simulator with a complex model (e.g., as we did for MTBD-CT(*κ*) models) than to derive the model likelihood (which we were only able to solve for BD-CT(1)). Using a simulation-based and likelihood-free machine learning approach [21] for future −CT model developments is hence a promising direction.

Contact-tracing is not the only mechanism leading to non-random sampling. Other mechanisms include community-based testing campaigns [28] and sampling policies varying over time and between locations. New models may need to be developed in the future to include and distinguish between these mechanisms. For instance, an index case and their potential contacts are more likely to belong to the same community; in this case, our cherry test would likely be positive, meaning that its results must be interpreted with caution as a possible indication of contact tracing, among other possible causes of non-random sampling.

Finally, what we presented here is a specific case of an epidemiological model. However, it might be useful in other contexts as well. For instance, the State Speciation and Extinction (SSE) family of models [8, 44, 45] in the macroevolutionary domain are mathematically very similar to the epidemiological MTBD models [46]. In macroevolution, a detection (or sequencing) of a new species might lead to faster detection (or sequencing) of closely related ones. This process could be modeled in the same way as contact tracing.

## Data Availability

All data produced are available online at https://github.com/evolbioinfo/bdct

## Acknowledgments

The authors would like to thank Dr Mathieu Moslonka-Lefebvre who initiated this project by proposing to study contact tracing. He worked on a different version of the CT model (which is not part of this manuscript) and on the initial version of the CT test. We would like to thank Dr Jakub Voznica for fruitful discussions on contact tracing mechanisms. We would also like to thank the HPC cluster team in Institut Pasteur for their support with computer simulations. O.G. was supported by the Paris Artificial Intelligence Research Institute (PRAIRIE, ANR-19-P3IA-0001).

## Supporting information

**S1 Appendix MTBD-CT tree simulator**. The appendix describes the tree simulator that we developed for generating sampled transmission trees under MTBD and MTBD-CT models.

**S2 Appendix. Calculation of a probability of a branch containing a hidden contact**. The appendix describes the approximation of the probability of having a branch in the tree that contains one or two hidden contacts.

**S3 Appendix. Tree likelihood calculation**. The appendix describes tree likelihood calculation case by case, accounting for all possible notifier-contact configurations.

