## Supplementary material for "Accounting for contact tracing in epidemiological birth-death models": S1 Appendix

### S1 Appendix: MTBD-CT tree simulator

Anna Zhukova, Olivier Gascuel

Our tree simulator generates sampled transmission trees under MTBD and MTBD-CT models (with $m$ states). It is Gillespie-based, generates state change, transmission and removal times, and only reconstructs the sampled parts of the tree to save memory and increase speed.

The simulator iterates through occurring events till either time or sampled tip number limit is reached. It keeps updating an array C of length $m$ containing counts of currently infected individuals (i.e., non-removed) in different states; an array S of length $m$ containing counts of sampled individuals in different states; as well as a mapping M between states and ids of currently infected individuals in those states, and a mapping N between states and ids of sampled individuals in those states. In the beginning ($t = 0$) the only infected individual corresponds to the root: $\mathrm{C}_k = 0$, $\mathrm{M}_k = \emptyset \ \forall k \neq \mathrm{state}(root)$; $\mathrm{C}_{\mathrm{state}(root)} = 1$, $\mathrm{M}_{\mathrm{state}(root)} = \{root\}$, and no individual is sampled: $\mathrm{S}_k = 0$, $\mathrm{N}_k = \emptyset$ $\forall 1 \leq k \leq m$.

At each iteration the algorithm (1) calculates the time of the next event; (2) chooses the type of this event and the the individual involved in it; (3) updates the counts and the mappings according to the event and records its time.

At step 1, to calculate the time of the next event, we (i) calculate the total rate $r$ as a sum of total state change, transmission and transition rates: $r = r^{(\mu)} + r^{(\lambda)} + r^{(\psi)}$, where $r^{(\mu)} = \sum_{k=1}^{m} \mathrm{C}_k \sum_{r=1}^{m} \mu_{kr}$; $r^{(\lambda)} = \sum_{k=1}^{m} \mathrm{C}_k \sum_{r=1}^{m} \lambda_{kr}$, and $r^{(\psi)} = \sum_{k=1}^{m} \mathrm{C}_k \psi_k$; (ii) draw $\Delta t$ from the exponential distribution with rate $r$; (iii) update the current time to $t + \Delta t$.

At step 2, to chose the type of the event, we draw a value $w$ (uniformly) from an interval $[0, r[$. This interval can be seen as composed of sub-intervals corresponding to each possible event, where the width of each sub-interval is defined by the corresponding rate and infected individual count, e.g., the sub-interval corresponding to a transmission from $k$ to $l$ has a width of $\lambda_{kl}\mathrm{C}_k$. The sub-interval in which $w$ is located defines the event type.

At step 3, we randomly draw an individual $i$ of type $k$ (selected at step 2) from $\mathrm{M}_k$, and proceed depending on the event type. If it is a state-change from $k$ to $l$, then we decrease $\mathrm{C}_k$ by one, increase $\mathrm{C}_l$ by one, remove $i$ from $\mathrm{M}_k$ and put $i$ into $\mathrm{M}_l$. If it is a transmission from $k$ to $l$, then we increase $\mathrm{C}_l$ by one, create a new id $j$ for the recipient and put $j$ into $\mathrm{M}_l$. If it is a removal, then we decrease $\mathrm{C}_k$ by one, remove $i$ from $\mathrm{M}_k$, and draw a value $p$ (uniformly) from an interval $[0, 1[$: if $p < \rho$, the individual gets sampled and we put $i$ into $\mathrm{N}_k$.

For transmission and sampling events we also record their time and ids of the involved individuals. These values are used to reconstruct the sampled parts of the tree once the simulation is finished.

For the -CT($\kappa$) version of this simulator, we add additional $m$ states for notified-contact versions of each state. $m + k$'s state-change rates are set to the $k$'s state-change rates : $\mu_{m+k,m+l} = \mu_{k,l} \ \forall 1 \leq k, l \leq m$, while the state-change rates between initial and notified states are set to zero: $\mu_{m+k,l} = \mu_{k,m+l} = 0$. Only the transmissions to non-notified states are allowed (and the ones from notified states are set to the corresponding non-notified rates): $\lambda_{m+k,l} = \lambda_{k,l}$, $\lambda_{m+k,l+k} = \lambda_{k,m+l} = 0$ $\forall 1 \leq k, l \leq m$. The removal rates of notified states are set to $\phi$: $\psi_{m+k} = \phi >> \psi_k$, while their sampling probabilities are set to 1: $\rho_{m+k} = 1 \ \forall 1 \leq k \leq m$. We keep a mapping of contacts P for each infectious individual (ordered from the last to the first), and update it for both the donor and the recipient at each transmission event. We also keep a mapping between each infectious individual and their state, and update it at each state-change event. At each removal event, if the removed individual $i$ gets sampled upon removal, we draw $\kappa$ values $p_l$ $1 \leq l \leq \kappa$ (uniformly) from an interval $[0, 1[$. If $p_l < \upsilon$, and if $i$ had at least $l$ contacts and the $l^{\text{th}}$-to-last contact ($j = \mathrm{P}_{il}$) is not yet sampled (not in $\mathrm{N}[\mathrm{state}(j)]$) and not yet notified ($\mathrm{state}(j) \leq m$), we change $j$'s state to $m + \mathrm{state}(j)$ as in a state-change event.

#### Code availability

The simulator is implemented in Python 3. It uses ETE 3 framework for tree manipulation [1] and NumPy package for array operations [2].

It is available as a command-line program and a Python 3 package via PyPi (treesimulator), and via Docker/Singularity (evolbioinfo/treesimulator). Its source code and the installation and usage documentation are available on GitHub at github.com/evolbioinfo/treesimulator.
