## Supplementary material for "Accounting for contact tracing in epidemiological birth-death models": S2 Appendix

### S2 Appendix: Mixed branch probability

Anna Zhukova, Olivier Gascuel

To model the cases when a contact B is not sampled, we need to calculate the probability of a **hidden contact subtree** $U_p(t, t_A)$, which started at time $t$ on a contact branch and evolved till time $T$ without any of its individuals being sampled, provided that the first of the contact's notifiers tried to notify them at time $t_A$ (B's branch in Fig. 2 (2a, 2c, 3a, 3c) in the main text). This probability consists of two parts: either the contact got notified at time $t_A$ but has not got sampled by the end of the sampling period $T$ (like in Fig. 2 (2a, 3a) in the main text), $U_p^{(1)}(t, t_A)$; or the contact got removed via the standard procedure before the time $t_A$ (like in Fig. 2(2c, 3c) in the main text), $U_p^{(2)}(t, t_A)$:

$$U_p(t, t_A) = U_p^{(1)}(t, t_A) + U_p^{(2)}(t, t_A). \tag{1}$$

The first part can be easily expressed via the previously defined equations: contact branch evolution before notification till time $t_A$ (if $t_A > t$) followed by contact branch evolution after notification till time $T$. Note that neither $p_{cb}^{(x)}(t, t_A)$ not $p_{ca}^{(x)}(t_A)$ include an event at the end of the branch, and in this case the contact branch does not end at $t_x = T$:

$$U_p^{(1)}(t, t_A) = \begin{cases} p_{cb}^{(A)}(t) p_{ca}^{(x)}(t_A) & \textit{if } t_A > t \\ p_{ca}^{(x)}(t) & \textit{if } t_A \leq t \end{cases}, \textit{ where } t_x = T \tag{2}$$

The second scenario is only possible if $t_A > t$. To calculate it, we would need to integrate over all possible contact removal times $t_B \in [t, t_A[$, provided that in the interval between $t$ and $t_B$ the contact might have transmitted to someone else (potentially multiple times), whose subtree(s) stayed unobserved till $T$. Note that $p_{cb}^{(A)}(t)$ describes a contact branch evolution between $t$ and $t_A$ with any number of hidden transmissions, no removal along it, and no event at the end of the branch. What we need instead is a removal with no sampling at time $t_B$. To approximate $U_p^{(2)}(t, t_A)$ we will divide $p_{cb}^{(A)}(t)$ by the probability of no removal during the time $(t_A - t)$ (i.e., $e^{-\psi(t_A - t)}$), multiply it by the probability of removal during that time $(1 - e^{-\psi(t_A - t)})$, and by the probability of not being sampled upon removal $(1 - \rho)$:

$$U_p^{(2)}(t, t_A) \approx \begin{cases} p_{cb}^{(A)}(t) \frac{1 - e^{-\psi(t_A - t)}}{e^{-\psi(t_A - t)}} (1 - \rho) & \textit{if } t_A > t \\ 0 & \textit{if } t_A \leq t \end{cases} \tag{3}$$

We now describe the probability of a **mixed branch** $p_{m(s_1, s_2)}^{(i)}(t, t_r)$ for branches that include a hidden contact subtree (AB-C branches in Fig. 2 (2) and A's tip branches in Fig. 2 (3) in the main text). $s_1$ here denotes the type of the top of the branch (e.g., between AB and BC in Fig. 2 (2), contact) and $s_2$ denotes the type of the bottom of the branch (e.g., between AB and A in Fig. 2 (3), notifier). Let us denote the first notification time of the hidden contact as $t_r$, and the time of the hidden contact subtree start as $t_h$, while as usual $t_i$ stands for the time at the branch end. For instance, in Fig. 2 (2a) $t_i = t_C$, $t_r = t_A$, $t_h = t_{BC}$; in Fig. 2 (2b) $t_i = t_r = t_C$, $t_h = t_{BC}$; in Fig. 2 (3c) $t_i = t_r = t_A$, $t_h = t_{AB}$ in the main text.

$p^{(i)}_{m(s_1,s_2)}(t,t_r)$ combines two elements: the probability of the top part of the branch $p^{(h)}_{top=s_1}(t,t_r)$, which finishes with a transmission to or from the hidden contact at time $t_h$ and the probability of the bottom part of the branch (without taking into account the event at its end) $p^{(i)}_{bottom=s_2}(t_h)$.

Note that since the branch contains a hidden contact, this contact must have at least one notifier (who is observed as all the notifiers). The notifier could correspond to a tip in the branch's supertree: For example, tip A who is part of the C's supertree in Fig. 2 (2a, 2c) in the main text. If the branch is external, the notifier could also correspond to its own tip: For example, such notifiers correspond to tip A in Fig. 2 (2b) and tip C in Fig. 2 (3b) in the main text. Having a notifier as the branch's tip implies that the bottom part of the branch corresponds to the notifier. Having a notifier in the supertree implies that the top part of the branch corresponds to the contact (already notified or not yet). Hence if the branch is internal, the top part can only correspond to the contact, while the bottom part can only correspond to a standard state (unnotified non-notifier), as the bottom part of such a branch corresponds to the contact's recipient (who could not be notified as the contact is unobserved and could not be a notifier as the branch is not external). Finally, if the branch is external, and the only contact's notifier corresponds to its tip, then there are two possibilities for its top part: (1a) either it fully corresponds to a standard state (unnotified non-notifier); or (1b) there is another contact's notifier in its supertree, and this other contact is also hidden along the top part of the same branch, hence the top part of the branch is contact-standard (see SFig. 1). If, on the other hand, the branch is external, and the contact's notifier(s) is(are) only found in its supertree, then there are two possibilities for its bottom part: (2a) either it fully corresponds to a standard state (unnotified non-notifier); or (2b) there is another contact's notifier at its tip, and this other contact is also hidden along the bottom part of the same branch, hence the bottom part of the branch is standard-notifier (see SFig. 1). Note that (1b) and (2b) represent the same case but from the point of view of two different hidden contacts (B and their notifier A for the former, and D and their notifier C for the latter in SFig. 1).

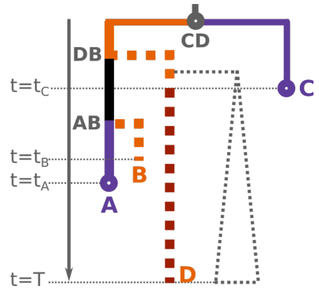


**SFig 1. An example of mixed tip branch with two hidden contact subtrees**: an individual C got sampled at time $t_C$ and notified their last contact D. D however, stayed unobserved as D did not get sampled by the end of the sampling period (time $T$). Before notification (at time $t_C$) D transmitted two times, the latter transmission leading to an unobserved recipient subtree (gray dotted triangle) and the former transmission (DB) leading to an observed recipient subtree (which includes a sampled individual A and a hidden individual B). The D's recipient A transmitted further (transmission AB). Once A got sampled at time $t_A$, they notified their last contact B, who however was already removed by then via the standard procedure without sampling (at time $t_B < t_A$). Hence the branch CD-A represents a mixed branch with states contact(CD-DB)-standard(DB-AB)-notifier(AB-A).

We will first describe the one-hidden-contact case $p^{(i)}_{m(s_1,s_2)}(t,t_r)$ (where $s_1 \in \{c,-\}$,

$s_2 \in \{n, -\}$ and $-$ stands for the standard (unnotified non-notifier) state), and then proceed with the two-hidden-contacts case $p^{(i)}_{m(c,-,n)}(t, t_r)$, where $t_r$ is the earliest notification time for the contact hidden at the top of the branch (e.g., $t_r = t_C$ in SFig. 1), the only notification time for the contact hidden at the bottom of the branch being $t_i$ (e.g., $t_i = t_A$ in SFig. 1).

For the bottom branch part:

$$p^{(i)}_{bottom=s_2}(t_h) = \begin{cases} p^{(i)}_n(t_h) & \textit{if } i \textit{ is a notifier tip } (s_2 = n), \\ p^{(i)}(t_h) & \textit{otherwise } (s_2 = -, \textit{ the standard state}) \end{cases} \tag{4}$$

We will approximate $p^{(h)}_{top=s_1}(t)$ using the branch evolution probability of the type corresponding to $s_1$ in the following way. For example, let us assume that $s_1 = -$ (standard branch). $p^{(h)}(t)$ expresses a probability of evolving along this branch between the times $t$ and $t_h$ with no or any number of hidden transmissions, without taking into account the event at the end of it. In our case there must be at least one hidden transmission: to/from the contact at time $t_h$ (where the contact tree stayed unsampled). If we remove the probability of no event during this time ($e^{-(\lambda+\psi)(t_h-t)}$) from $p^{(h)}(t)$, we obtain a probability of a branch with at least one hidden transmission somewhere between $t$ and $t_h$ (the probability of the corresponding hidden tree being included). $p^{(h)}_{top=-}(t)$ expresses almost the same thing, with the difference being that the last hidden transmission must have happened at time $t_h$ and correspond to the hidden contact. We will hence approximate $p^{(h)}_{top=-}(t)$ as follows:

$$p^{(h)}_{top=-}(t, t_r) \approx p^{(h)}(t) - e^{-(\lambda+\psi)(t_h-t)} \tag{5}$$

We will treat the case when the branch is in the contact state ($s_1 = c$) in a similar way:

$$p^{(h)}_{top=c}(t, t_r) \approx \begin{cases} p^{(h)}_{cb}(t) - e^{-(\lambda+\psi)(t_h-t)} & \textit{if } t_h \le t_r \\ p^{(r)}_{cb}(t)\left(p^{(h)}_{ca}(t_r) - e^{-(\lambda+\phi)(t_h-t_r)}\right) & \textit{if } t \le t_r \le t_h \\ p^{(h)}_{ca}(t) - e^{-(\lambda+\phi)(t_h-t)} & \textit{if } t > t_r \end{cases}, \tag{6}$$

where $t_r$ is the first notification time of the hidden contact.

Finally, we will account for the fact that the hidden tree starting at time $t_h$ corresponds to the hidden contact by dividing the expression by $U(t_h)$ (a standard hidden tree) and multiplying by $U_p(t_h, t_r)$ (contact hidden tree).

Putting everything together, and approximating the time of the start of the hidden contact tree $t_h$ with the middle of the branch, we obtain:

$$p^{(i)}_{m(s_1,s_2)}(t, t_r) = p^{(h)}_{top=s_1}(t, t_r)\frac{U_p(t_h, t_r)}{U(t_h)}p^{(i)}_{bottom=s_2}(t_h), \tag{7}$$

where $t_h \approx t + \frac{(t_i-t)}{2}$ is an approximation of the time of the hidden contact subtree start, $t_r$ is the earliest notification time of the hidden contact.

Finally, we approximate the mixed case with two hidden contacts using similar principals:

$$p^{(i)}_{m(c,-,n)}(t, t_r) \approx p^{(h1)}_{top=c}(t, t_r)\frac{U_p(t_{h1}, t_r)}{U(t_{h1})}p^{(h2)}_{top=-}(t_{h1}, t_i)\frac{U_p(t_{h2}, t_i)}{U(t_{h2})}p^{(i)}_n(t_{h2}), \tag{8}$$

where $t_{h1} \approx t + \frac{1}{3}(t_i - t)$, $t_{h2} \approx t + \frac{2}{3}(t_i - t)$, $t_r$ is the earliest notification time of the contact hidden at the top of the branch.
