## Supplementary material for "Accounting for contact tracing in epidemiological birth-death models": S3 Appendix

### S3 Appendix: Tree likelihood calculation under BD-CT(1) model

Anna Zhukova, Olivier Gascuel

#### Case 1: $i$ is a tip

For a tip $i$ with a parent node $j$ the subtree likelihood density values can be calculated as:

$$\begin{aligned} l^{(\bar{n},\bar{n})}(i) &= p^{(i)}(t_j)\psi\rho(1-\upsilon) \text{ i.e., } i \text{ was not notified and did not notify} \\ &\quad \text{(standard branch evolution followed by sampling and no notification)} \\ &\quad + p^{(i)}_{m(-,n)}(t_j,t_i)\psi\rho\upsilon \text{ i.e., } i \text{ was not notified but notified their contact who is hidden} \\ &\quad \text{(mixed standard-notifier branch evolution followed by sampling and notification)} \end{aligned} \tag{1}$$

$$\begin{aligned} l^{(\bar{n},n)}(i) &= p^{(i)}_{n}(t_j)\psi\rho\upsilon \text{ i.e., } i \text{ was not notified but notified their last contact (observed)} \\ &\quad \text{(notifier branch evolution followed by sampling and notification)} \end{aligned} \tag{2}$$

$$l^{(n,\bar{n})}(i,r) = \begin{cases} p^{(i)}_{ca}(t_j)\phi(1-\upsilon) \text{ if } i \text{ got notified at time } t_r < t_j \text{ and did not notify} \\ \text{(notified contact branch evolution then notification-provoked sampling and no notification)} \\ \quad + p^{(i)}_{m(ca,-)}(t_j,t_r)\psi\rho(1-\upsilon) \text{ i.e., branch } i \text{ contains a hidden contact notified by } r \\ \qquad \text{at } t_r < t_j, \text{ } i \text{ did not notify} \\ \quad \text{(mixed contact-standard branch evolution followed by sampling and no notification)} \\ \quad + p^{(i)}_{m(ca,n)}(t_j,t_r)\psi\rho\upsilon \text{ i.e., branch } i \text{ contains a hidden contact notified both by } r \text{ and } i \\ \quad \text{(mixed contact-notifier branch evolution followed by sampling and notification)} \\ \quad + p^{(i)}_{m(ca,-,n)}(t_j,t_r)\psi\rho\upsilon \text{ i.e., branch } i \text{ contains a hidden contact notified by } r \\ \qquad \text{at } t_r < t_j, \text{ } i \text{ notified another hidden contact} \\ \quad \text{(mixed contact-standard-notifier branch evolution then sampling and notification)} \\ \\ p^{(i)}_{cb}(t_j)\psi\rho(1-\upsilon) \text{ if } i \text{ got notified at time } t_r > t_i \text{ and did not notify} \\ \text{(contact-before-notification branch evolution then standard sampling and no notification)} \\ \quad + p^{(i)}_{m(cb,-)}(t_j,t_r)\psi\rho(1-\upsilon) \text{ i.e., branch } i \text{ contains a hidden contact notified by } r \\ \qquad \text{at } t_r > t_i, \text{ } i \text{ did not notify} \\ \quad \text{(mixed contact-standard branch evolution followed by sampling and no notification)} \\ \quad + p^{(i)}_{m(cb,n)}(t_j,t_i)\psi\rho\upsilon \text{ i.e., branch } i \text{ contains a hidden contact notified both by } r \text{ and } i \\ \quad \text{(mixed contact-notifier branch evolution followed by sampling and notification)} \\ \quad + p^{(i)}_{m(cb,-,n)}(t_j,t_r)\psi\rho\upsilon \text{ i.e., branch } i \text{ contains a hidden contact notified by } r \text{ at } t_r > t_i, \\ \qquad i \text{ notified another hidden contact} \\ \quad \text{(mixed contact-standard-notifier branch evolution then sampling and notification)} \\ \\ p^{(r)}_{cb}(t_j)p^{(i)}_{ca}(t_r)\phi(1-\upsilon) \text{ if } i \text{ got notified at time } t_j \le t_r \le t_i \text{ and did not notify} \\ \text{(contact-before-notification branch evolution followed by} \\ \text{ notified contact branch evolution, notification-provoked sampling and no notification)} \\ \quad + p^{(i)}_{m(c,-)}(t_j,t_r)\psi\rho(1-\upsilon) \text{ i.e., branch } i \text{ contains a hidden contact notified by } r \\ \qquad \text{at } t_j \le t_r \le t_i, \text{ } i \text{ did not notify} \\ \quad \text{(mixed contact-standard branch evolution followed by sampling and no notification)} \\ \quad + p^{(i)}_{m(c,n)}(t_j,t_r)\psi\rho\upsilon \text{ i.e., branch } i \text{ contains a hidden contact notified both by } r \text{ and } i \\ \quad \text{(mixed contact-notifier branch evolution followed by sampling and notification)} \\ \quad + p^{(i)}_{m(c,-,n)}(t_j,t_r)\psi\rho\upsilon \text{ i.e., branch } i \text{ contains a hidden contact notified by } r \\ \qquad \text{at } t_j \le t_r \le t_i, \text{ } i \text{ notified another hidden contact} \\ \quad \text{(mixed contact-standard-notifier branch evolution then sampling and notification)} \end{cases} \tag{3}$$

$$
l^{(n,n)}(i,r) = \begin{cases} p_b^{(i)}(t_j)\phi\upsilon \text{ if } i \text{ got notified at time } t_r < t_j \text{ and notified their last contact} \\ \text{(notified notifier branch evolution then notification-provoked sampling and notification)} \\ \\ p_n^{(i)}(t_j)\psi\rho\upsilon \text{ if } i \text{ got notified at time } t_r > t_i \text{ and notified their last contact} \\ \text{(notifier branch evolution followed by standard sampling and notification)} \\ \\ p_n^{(r)}(t_j)p_b^{(i)}(t_r)\phi\upsilon \text{ if } i \text{ got notified at time } t_j \le t_r \le t_i \text{ and notified} \\ \text{(notifier branch evolution followed by notified notifier branch evolution,} \\ \text{ notification-provoked sampling and notification)} \end{cases} \tag{4}
$$

#### Case 2: $i$ is an internal node

Let us now consider an internal node $i$ with two child nodes, $i_0$ and $i_1$.

#### Case 2.1: $i$ is an unnotified internal node

We will start with the configuration where the $i$'s branch corresponds to an unnotified individual. Hence the $i$'s branch evolution is represented by $p^{(i)}(t_j)$ with a transmission event at the end of it, where each of the child branches can correspond to the donor (probability density of $2\lambda$). We will consider three possibilities: (2.1.1) when both $i_0$ and $i_1$ are internal nodes; (2.1.2) when both $i_0$ and $i_1$ are tips; and (2.1.3) when one of them is an internal node and the other one is a tip.

#### Case 2.1.1: $i$ is an unnotified internal node with internal-node children $i_0$ and $i_1$

First, let's assume that $i_0$ and $i_1$ are internal. Then none of them can be a notifier (since notifiers correspond to tips), and hence none of them is notified (as $i$ is also unnotified):

$$
l^{(\bar{n})}(i) = p^{(i)}(t_j)2\lambda l^{(\bar{n})}(i_0)l^{(\bar{n})}(i_1) \tag{5}
$$

#### Case 2.1.2: $i$ is an unnotified internal node with tip children $i_0$ and $i_1$

Secondly, let's assume that $i_0$ and $i_1$ are tips. Then each of them could have notified the other one:

$$
\begin{aligned} l^{(\bar{n})}(i) = p^{(i)}(t_j)2\lambda \cdot \Big( & l^{(\bar{n},\bar{n})}(i_0)l^{(\bar{n},\bar{n})}(i_1) \leftarrow \text{neigher } i_0 \text{ nor } i_1 \text{ notified} \\ & + l^{(\bar{n},n)}(i_0)l^{(n,\bar{n})}(i_1,i_0) \leftarrow i_0 \text{ notified } i_1 \text{ and } i_1 \text{ did not notify} \\ & + l^{(n,\bar{n})}(i_0,i_1)l^{(\bar{n},n)}(i_1) \leftarrow i_0 \text{ did not notify, but } i_1 \text{ notified } i_0 \\ & + l^{(n,n)}(i_0,i_1)l^{(n,n)}(i_1,i_0) \Big) \leftarrow i_1 \text{ and } i_0 \text{ notified each other} \end{aligned} \tag{6}
$$

#### Case 2.1.3: $i$ is an unnotified internal node whose children are a tip and an internal node

Lastly, let's assume that $i_0$ is a tip and $i_1$ is internal (the scenario where $i_1$ is a tip and $i_0$ is internal can be obtained by swapping the labels). Then $i_0$ could have notified $i_1$:

$$
\begin{aligned} l^{(\bar{n})}(i) = p^{(i)}(t_j)2\lambda \cdot \Big( & l^{(\bar{n},\bar{n})}(i_0)l^{(\bar{n})}(i_1) \leftarrow i_0 \text{ did not notify } i_1 \\ & + l^{(\bar{n},n)}(i_0)l^{(n)}(i_1,i_0) \Big) \leftarrow i_0 \text{ notified } i_1 \end{aligned} \tag{7}
$$

#### Case 2.2: $i$'s branch is internal and (at least initially) corresponds to an (eventually notified) contact

In the other case the internal node $i$'s branch starts as a contact who will eventually get notified by a tip $r$. There are two possibilities: either the node $i$ corresponds to the contact or the contact is hidden along the $i$'s branch and $i$ corresponds to someone within $i$'s recipient subtree (e.g., if $i = C$ in Fig. **??**(2a)). In the first case, the $i$'s branch evolution is represented by the contact branch evolution $p_c^{(i)}(t_j, t_r)$:

$$p_c^{(i)}(t_j, t_r) = \begin{cases} p_{ca}^{(i)}(t_j) & \textit{if } t_r < t_j, \\ p_{cb}^{(i)}(t_j) & \textit{if } t_r > t_i, \\ p_{cb}^{(r)}(t_j) p_{ca}^{(i)}(t_r) & \textit{if } t_j \le t_r \le t_i. \end{cases} \tag{8}$$

This branch has a transmission event at the end of it, where the donor is known (probability density of $\lambda$): The donor corresponds to the subtree containing the tip representing the sampling of the individual $i$.

In the second case, the $i$'s branch evolution is represented by $p_{m(c,-)}^{(i)}(t_j, t_r)$ with a transmission event at the end of it, where the donor is unknown (probability density of $2\lambda$): any of the subtrees can be the donor.

Again, we will consider three possibilities for $i$'s child nodes: (2.2.1) when both $i_0$ and $i_1$ are internal nodes; (2.2.2) when both $i_0$ and $i_1$ are tips; and (2.2.3) when one of them is an internal node and the other one is a tip.

#### Case 2.2.1: $i$'s branch is internal and (at least initially) corresponds to a contact with internal-node children $i_0$ and $i_1$

First, let's assume that $i_0$ and $i_1$ are internal. Then none of them can be a notifier (since notifiers correspond to tips), and hence either exactly one of them is notified by $r$, or the contact is hidden:

$$\begin{aligned} l^{(n)}(i, r) = & p_c^{(i)}(t_j, t_r)\lambda \cdot \Big( l^{(n)}(i_0, r) l^{(\bar{n})}(i_1) \leftarrow i_0 \textit{ is notified by } r \\ & \qquad + l^{(\bar{n})}(i_0) l^{(n)}(i_1, r) \Big) \leftarrow i_1 \textit{ is notified by } r \\ & + p_{m(c,-)}^{(i)}(t_j, t_r) 2\lambda l^{(\bar{n})}(i_0) l^{(\bar{n})}(i_1) \leftarrow \textit{hidden contact} \end{aligned} \tag{9}$$

#### Case 2.2.2: $i$'s branch is internal and (at least initially) corresponds to a contact with tip children $i_0$ and $i_1$

Secondly, let's assume that $i_0$ and $i_1$ are tips. Then each of them could have notified the other one (in addition to one of them potentially being notified by $r$):

$$
\begin{aligned}
l^{(n)}(i,r) =& p_c^{(i)}(t_j,t_r)\lambda \cdot \Big( \, l^{(n,\bar{n})}(i_0, \text{first}(r,i_1)) l^{(\bar{n},n)}(i_1) \leftarrow \textit{r and } i_1 \textit{ notified } i_0,\ i_0 \textit{ did not notify} \\
&+ l^{(n,n)}(i_0, \text{first}(r,i_1)) l^{(n,n)}(i_1, i_0) \leftarrow \textit{r and } i_1 \textit{ notified } i_0,\ i_0 \textit{ notified } i_1 \\
&+ l^{(\bar{n},n)}(i_0) l^{(n,\bar{n})}(i_1, \text{first}(r,i_0)) \leftarrow \textit{r and } i_0 \textit{ notified } i_1,\ i_1 \textit{ did not notify} \\
&+ l^{(n,n)}(i_0, i_1) l^{(n,n)}(i_1, \text{first}(r,i_0)) \leftarrow \textit{r and } i_0 \textit{ notified } i_1,\ i_1 \textit{ notified } i_0 \\
&+ l^{(n,n)}(i_0, r) l^{(n,\bar{n})}(i_1, i_0) \leftarrow \textit{r notified } i_0,\ i_0 \textit{ notified } i_1,\ i_1 \textit{ did not notify} \\
&+ l^{(n,\bar{n})}(i_0, i_1) l^{(n,n)}(i_1, r) \leftarrow \textit{r notified } i_1,\ i_1 \textit{ notified } i_0,\ i_0 \textit{ did not notify} \\
&+ l^{(n,\bar{n})}(i_0, r) l^{(\bar{n},\bar{n})}(i_1) \leftarrow \textit{r notified } i_0,\ i_0 \textit{ and } i_1 \textit{ did not notify} \\
&+ l^{(\bar{n},\bar{n})}(i_0) l^{(n,\bar{n})}(i_1, r) \, \Big) \leftarrow \textit{r notified } i_1,\ i_0 \textit{ and } i_1 \textit{ did not notify} \\
&+ p_{m(c,-)}^{(i)}(t_j,t_r) 2\lambda \cdot \Big( \, l^{(n,\bar{n})}(i_0, i_1) l^{(\bar{n},n)}(i_1) \leftarrow \textit{r notified a hidden contact,} \\
&\qquad i_1 \textit{ notified } i_0,\ i_0 \textit{ did not notify} \\
&+ l^{(n,n)}(i_0, i_1) l^{(n,n)}(i_1, i_0) \leftarrow \textit{r notified a hidden contact,} \\
&\qquad i_1 \textit{ notified } i_0,\ i_0 \textit{ notified } i_1 \\
&+ l^{(\bar{n},n)}(i_0) l^{(n,\bar{n})}(i_1, i_0) \leftarrow \textit{r notified a hidden contact,} \\
&\qquad i_0 \textit{ notified } i_1,\ i_1 \textit{ did not notify} \\
&+ l^{(\bar{n},\bar{n})}(i_0) l^{(\bar{n},\bar{n})}(i_1) \, \Big) \leftarrow \textit{r notified a hidden contact,} \\
&\qquad i_0 \textit{ and } i_1 \textit{ did not notify}
\end{aligned}
\tag{10}
$$

where $\text{first}(a,b) = \begin{cases} a \ \textit{if} \ t_a \le t_b \\ b \ \textit{if} \ t_a > t_b \end{cases}$ .

**Case 2.2.3: $i$'s branch is internal and (at least initially) corresponds to a contact, $i$'s children are a tip and an internal node**

Lastly, let's assume that $i_0$ is a tip and $i_1$ is internal (the scenario where $i_1$ is a tip and $i_0$ is internal can be obtained by swapping the labels). Then $i_0$ could have notified $i_1$ (in addition to one of them being potentially notified by $r$):

$$
\begin{aligned}
l^{(n)}(i,r) =& p_c^{(i)}(t_j,t_r)\lambda \cdot \Big( \, l^{(n,n)}(i_0, r) l^{(n)}(i_1, i_0) \leftarrow \textit{r notified } i_0,\ i_0 \textit{ notified } i_1 \\
&+ l^{(n,\bar{n})}(i_0, r) l^{(\bar{n})}(i_1) \leftarrow \textit{r notified } i_0,\ i_0 \textit{ did not notify} \\
&+ l^{(\bar{n},n)}(i_0) l^{(n)}(i_1, \text{first}(r,i_0)) \leftarrow \textit{both r and } i_0 \textit{ notified } i_1 \\
&+ l^{(\bar{n},\bar{n})}(i_0) l^{(n)}(i_1, r) \, \Big) \leftarrow \textit{r notified } i_1,\ i_0 \textit{ did not notify} \\
&+ p_{m(c,-)}^{(i)}(t_j,t_r) 2\lambda \cdot \Big( \, l^{(\bar{n},n)}(i_0) l^{(n)}(i_1, i_0) \leftarrow \textit{r notified a hidden contact,} \\
&\qquad i_0 \textit{ notified } i_1 \\
&+ l^{(\bar{n},\bar{n})}(i_0) l^{(\bar{n})}(i_1) \, \Big) \leftarrow \textit{r notified a hidden contact,} \\
&\qquad i_0 \textit{ did not notify}
\end{aligned}
\tag{11}
$$

where $\text{first}(a,b) = \begin{cases} a \ \textit{if} \ t_a \le t_b \\ b \ \textit{if} \ t_a > t_b \end{cases}$ .

**Tree likelihood calculation with a pruning algorithm and its time complexity**

Note that $l^{(n)}(i,r)$, $l^{(n,n)}(i,r)$ and $l^{(n,\bar{n})}(i,r)$ depend on the notifier $r$ and the notification time $t_r$. Note that for a node $i$ any tip $r$ that is sister to any internal node on the path between $i$ and the tree root could be its potential notifier. During the tree likelihood calculation we first annotate each node $i$ with a set of potential notifiers $N_i$. For that we first add tips' to their parent nodes' potential notifier sets. We then perform a preorder tree traversal, where for every visited internal node $i$ we add its potential notifiers to its child nodes $i_0$ and $i_1$: $N_{i_0} := N_{i_0} \cup N_i - \{i_0\}$, $N_{i_1} := N_{i_1} \cup N_i - \{i_1\}$ (removing the child itself, as if it is a tip it will be found among its parent's potential notifiers). This preprocessing is only performed once, takes time O(N) where N is the number of tips in the tree and is negligible with respect to the rest of the parameter inference procedure.

Then we perform a postorder tree traversal, where for each node $i$ we calculate $l^{(\bar{n},\bar{n})}(i)$, $l^{(\bar{n},n)}(i)$, $l^{(n,\bar{n})}(i,r)$ and $l^{(n,n)}(i,r)$ $\forall r \in N_i$ if it is a tip, or $l^{(\bar{n})}(i)$ and $l^{(n)}(i,r)$ $\forall r \in N_i$ if it is internal. This part takes O(R + N) time, where O(N) accounts for calculation of $l^{(\bar{n})}(i)$ for all the $N-1$ internal tree nodes and of $l^{(\bar{n},\bar{n})}(i)$ and $l^{(\bar{n},n)}(i)$ for $N$ tips, while R is the sum of numbers of potential notifiers over all tree nodes and O(R) accounts for calculation of $l^{(n)}(i,r)$, $l^{(n,\bar{n})}(i,r)$ and $l^{(n,n)}(i,r)$ $\forall i$ $\forall r$.

For the best case (a perfectly balanced tree, where each tip is in a cherry), the number of potential notifiers is one for the tips (i.e., the sister tip) and zero for the internal nodes. Hence $R_{balanced} = N$ and the likelihood calculation time is linear: O(N). The worst case is represented by a caterpillar tree (where each internal node has a tip child). Each internal node has $k$ potential notifiers (tips in its supertree), where $k$ is the node's depth in the tree, the root's depth being 0 and the deepest internal node's depth being $N-2$. Moreover each tip has as many notifiers as its parent node, apart from the two deepest tips who have $N-1$ notifiers each instead of $N-2$ as they are in a cherry and hence could have notified each other. This hence sums up to $R_{caterpillar} = 2\big(1 + 2 + \ldots + (N-3)\big) + 2(N-1) = N^2 - 2N + 2$ and the time is hence quadratic: $O(N^2)$. On our BD-CT(1) simulated dataset of 100 trees with N=500-1 000 tips each, $R \in [2N, 38N]$ with a median at $7N$, suggesting a likelihood calculation time close to the linear one.
